# Genetic regulation of *TERT* splicing contributes to reduced or elevated cancer risk by altering cellular longevity and replicative potential

**DOI:** 10.1101/2024.11.04.24316722

**Authors:** Oscar Florez-Vargas, Michelle Ho, Maxwell Hogshead, Chia-Han Lee, Brenen W Papenberg, Kaitlin Forsythe, Kristine Jones, Wen Luo, Kedest Teshome, Cornelis Blauwendraat, Kimberly J Billingsley, Mikhail Kolmogorov, Melissa Meredith, Benedict Paten, Raj Chari, Chi Zhang, John S. Schneekloth, Mitchell J Machiela, Stephen J Chanock, Shahinaz Gadalla, Sharon A Savage, Sam M Mbulaiteye, Ludmila Prokunina-Olsson

**Author notes:** Corresponding author: Ludmila Prokunina-Olsson, PhD Laboratory of Translational Genomics, Division of Cancer Epidemiology and Genetics, National Cancer Institute, 9615 Medical Center Dr, Rockville, MD, 20850, USA.

## Abstract

The chromosome 5p15.33 region, which encodes telomerase reverse transcriptase (TERT), harbors multiple germline variants identified by genome-wide association studies (GWAS) as risk for some cancers but protective for others. We characterized a variable number tandem repeat within *TERT* intron 6 (VNTR6-1, 38-bp repeat unit) and observed a strong association between VNTR6-1 alleles (Short: 24-27 repeats, Long: 40.5-66.5 repeats) and GWAS signals within *TERT* intron 4. Specifically, VNTR6-1 fully explained the GWAS signals for rs2242652 and partially for rs10069690. VNTR6-1, rs10069690 and their haplotypes were associated with multi-cancer risk and age-related telomere shortening. Both variants reduce *TERT* expression through alternative splicing and nonsense-mediated decay: rs10069690-T increases intron 4 retention and VNTR6-1-Long expands a polymorphic G quadruplex (G4, 35-113 copies) within intron 6. Treatment with G4-stabilizing ligands decreased the fraction of the functional telomerase-encoding *TERT* full-length isoform, whereas CRISPR/Cas9 deletion of VNTR6-1 increased this fraction and apoptosis while reducing cell proliferation. Thus, VNTR6-1 and rs10069690 regulate the expression and splicing of *TERT* transcripts encoding both functional and nonfunctional telomerase. Altered TERT isoform ratios might modulate cellular longevity and replicative potential at homeostasis and in response to environmental factors, thus selectively contributing to the reduced or elevated cancer risk conferred by this locus.

## INTRODUCTION

*TERT* encodes the catalytic subunit of telomerase, a reverse transcriptase that extends telomeric repeats at chromosome ends, ensuring the maintenance of telomere length, genome integrity, and cell proliferation^1^. Telomere dysfunction has been implicated in many human diseases^2^. At least ten independent pleiotropic multi-cancer GWAS signals within the ∼100 kb genomic region on chromosome 5p15.33 harboring *TERT* and *CLPTM1L* have been associated with cancer risk or protection^3–6^. The associated variants might be causal or tag some known or yet unknown functional polymorphisms. Identifying these variants and the mechanisms underlying their associations may improve the understanding of the etiology and biological mechanisms of these cancers, leading to optimized cancer risk prediction, prevention, and treatment.

Several variable number tandem repeats (VNTRs) have been reported within the 5p15.33 region^7,8^ but only minimally characterized due to their high variability, complexity, and length of genomic fragments extended by repeat copies. Recent advances in long-read genome sequencing and assembly^9^ have closed many genomic gaps and facilitated the discovery and exploration of complex regions, such as VNTRs, for which copy numbers analysis using short-read sequencing or PCR-based methods is challenging. Recent examples^10^ have shown that VNTRs might account for or contribute to GWAS signals for cancer and other human traits, expanding the list of previously unknown functional genetic variants to be explored.

We hypothesized that VNTRs might be responsible for some of the reported GWAS signals within the *TERT* region. Here, we explored two VNTRs within *TERT* intron 6 in relation to the cancer-related GWAS signals reported in this region. Among those signals, a strong association was observed only between VNTR6-1 and two single nucleotide polymorphisms (SNPs) - rs2242652 and rs10069690 - within *TERT* intron 4. Specifically, VNTR6-1 Long alleles (40.5-66.5 repeats) in contrast to Short alleles (24-27 repeats) were preferentially linked with rs2242652-A and rs10069690-T alleles, both of which are associated with a reduced risk of bladder^6^ and prostate cancer^11^ but an elevated risk of glioma^12^, breast cancer^13,14^ and ovarian cancer^15^. We present a comprehensive genetic and functional analysis of VNTR6-1 and GWAS signals within this region. These results provide new insights into the etiology and genetic susceptibility of multiple cancers and telomere biology.

## RESULTS

### VNTR6-1 is linked with GWAS leads rs2242652 and rs10069690

We explored two previously reported but only minimally characterized VNTRs^7^,8 within t*he T*ERT intron 6 in relation to all cancer-related GWAS signals within the multi-cancer 5p15.33 region^3^-6. For this, we analyzed 452 phased long-read genome assemblies from 226 controls of diverse ancestries generated by the Human Pangenome Reference Consortium (HPRC)9 and the Center for Alzheimer’s and Related Dementias (CARD)^16^. The strongest associations were detected for VNTR6-1 (38-bp repeat unit, range 24-66.5 copies in the assemblies). Specifically, more VNTR6-1 copies were detected in assemblies with alleles of *TERT* intron 4 SNPs - rs2242652-A (p=5.93E-19) and rs10069690-T (p=5.40E-11) compared with the alternative alleles at these SNPs (**Figure 1a, b, Figure S1, Table S1**). In contrast, the copies of VNTR6-2 (36-bp repeat unit, range 8-155 copies in the assemblies) were only moderately associated with rs2242652-A allele (p=7.66-04) but not with rs10069690 (p=0.84, **Figure 1c**), or other GWAS signals (**Table S1**). Thus, we focused on VNTR6-1 as a potential functional proxy for GWAS leads rs2242652 and rs10069690.

**Figure 1.**
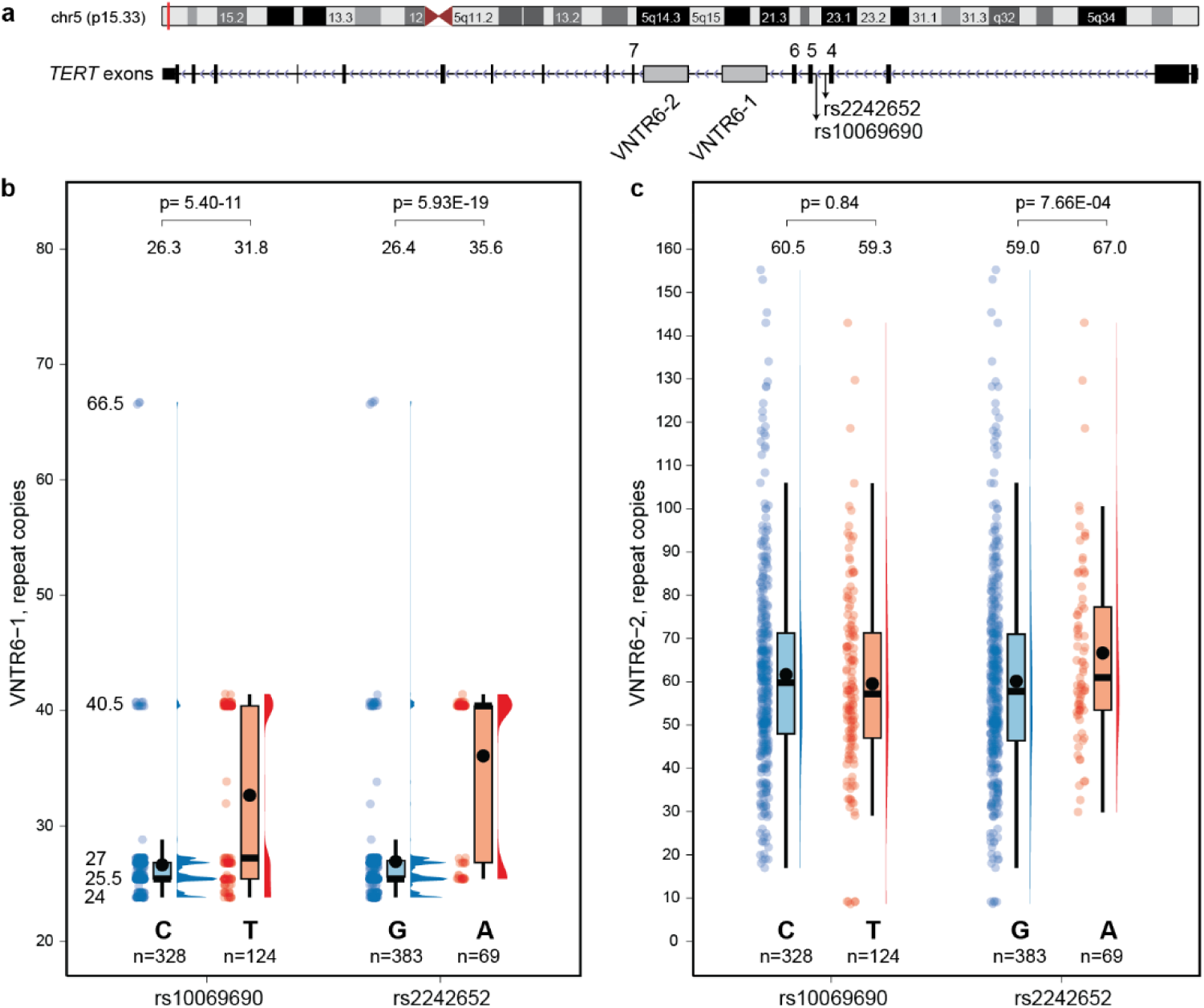
Analysis of VNTR6-1 and VNTR6-2 within *TERT* intron 6 in relation to GWAS leads rs10069690 and rs2242652. **a**, The chr 5p15.33 genomic region with GWAS leads rs10069690 and rs2242652 within *TERT* intron 4 and VNTRs within intron 6. **b**, Distribution of repeat copies of VNTR6-1 (38-bp repeat unit) and **c,** VNTR6-2 (36-bp repeat unit) in 452 phased long-read genomic assemblies from 226 controls of diverse ancestries. The dots represent repeat copies for each chromosome assembly; box plots show the overall and interquartile range, median (horizontal black line), and mean (black dot, with values above corresponding plots). Half-violin plots show the density distribution of the data. Five VNTR6-1 alleles—24, 25.5, 27, and 40.5 repeat copies—were observed above the 5% frequency threshold and accounted for 90.04% of all alleles in the set; VNTR6-2 alleles were scattered between 8 and 155 repeat copies, all under the 5% threshold. P values were calculated for unpaired two-sample Wilcoxon–Mann–Whitney tests comparing the number of repeat copies between the genotype groups.

We performed targeted PacBio sequencing of the VNTR6-1 amplicon (2126-3750 bp) in various samples (**Table S2**). This analysis confirmed concordance in repeat scoring in targeted vs. whole-genome long-read sequencing of 5 HPRC controls with available long-read assemblies^9^, in targeted sequencing of 5 pairs of tumor vs. tumor-adjacent normal bladder tissues, as well as Mendelian segregation in HapMap samples from 30 European (Central European from Utah, CEU) and 30 African (Yoruba, YRI) family trios. Two SNPs, rs56345976 and rs33961405 SNPs, were covered by the PacBio amplicon and their genotypes perfectly matched those determined by long-read genome assemblies and our TaqMan genotyping confirming that VNTR6-1 is a stable germline polymorphism that can be confidently genotyped by long-read sequencing (**Table S2**). Despite its reliability, long-read sequencing remains an expensive, laborious, and low-throughput method that requires a significant amount of high-quality DNA. The availability of more convenient approaches to analyze VNTR6-1 would facilitate its testing in association studies.

We noted that both in assemblies and in HapMap samples with targeted PacBio sequencing data (**Figure 1**, **Table S2**), the VNTR6-1 alleles clustered in 2 main groups. In HapMap samples, these groups comprised Short alleles (CEU: 25.8±1.0 copies, 83.3% and YRI: 27.0 ±2.0 copies, 80%) and Long alleles (CEU: 16.7%, 40.5 ±0 copies, and YRI: 43.75±8.8 copies, 20%). The Long alleles included an uncommon allele detected in YRI only: 66.5±0 copies, 2.5% (**Table S2**). Hypothesizing that short-read sequencing might help with VNTR analysis, we examined the genomic depth profiles of aligned short reads generated by whole-genome sequencing in all 3,201 individuals of diverse ancestries from the 1000 Genomes Project (1000G). The reference human genome has the Short VNTR6-1 allele (27 copies) and in 1000G samples carrying only the Short alleles (24-27 copies based on HPRC assemblies or targeted PacBio), the genomic profiles were flat. In contrast, noticeable read pileups were observed in 1000G samples with at least one copy of the Long allele (40.5 or 66.5 copies); however, the Long alleles and their heterozygous or homozygous state could not be distinguished based on the genomic profiles (**Figure S2**). We applied machine-learning methods to evaluate short-read profiles and classify all the 1000G samples as carriers of at least one copy of the VNTR6-1-Long allele (Long/any genotype) vs. the Short/Short genotype (**Table S3**). By treating the classification based on genomic profiles as true genotypes, we performed random forest analysis of the 1000G samples using all SNPs within the 400 kb genomic region (GRCh38 chr5:1,100,000-1,500,000) derived from short-read whole-genome sequencing and identified two SNPs, rs56345976 and rs33961405, that most effectively predicted VNTR6-1 groups in all populations despite differences in LD profiles (**Figure S3, Table S4, Table S5**). Although these SNPs were not sufficiently informative on their own, the rs56345976-A/rs33961405-G haplotype separated the carriers with VNTR6-1-Long alleles (40.5 or 66.5 copies) from the carriers of the VNTR6-1-Short/Short genotypes based on three other haplotypes (AA, GG, and GA) (**Figure S4, Table S3**).

The classification of all 1000G samples into groups with VNTR6-1 Short/Short and Long/any genotypes based on the rs56345976/rs33961405 haplotypes matched the scoring based on genomic profiles, with an area under the curve (AUC) of 0.98 (**Figure S4**). The VNTR6-1 scoring based on rs56345976/rs33961405 haplotypes was concordant with the repeat sizes determined by assemblies and targeted PacBio sequencing (**Figure S5, Table S6**). To test whether the VNTR6-1 can be confidently imputed, we created a custom reference panel of the region (400 kb) in all 3,201 samples from the 1000G dataset. VNTR6-1 was incorporated into this panel as a biallelic marker with Short/Long alleles determined based on phased rs56345976/rs33961405 haplotypes (**Table S3**). We randomly split the 1000G dataset into two equal groups and used the first group as a reference for imputing VNTR6-1 in the second group. The imputation showed a 99.3% concordance with the predetermined VNTR6-1 genotypes, further supporting the robustness of the VNTR6-1 allele assignment (Short/Long) based on rs56345976/rs33961405 haplotypes. These results demonstrated that VNTR6-1 could be used as a germline biallelic marker and confidently imputed into datasets with good coverage of the region through whole-genome sequencing or array genotyping and imputation. Because rs56345976/rs33961405 can be genotyped individually by targeted assays (such as by TaqMan assays), the VNTR6-1 genotypes can be inferred in any samples, even without genome-wide sequencing or genotyping data. In a subset of 1000G samples from Europeans (1000G-EUR), the VNTR6-1-Long allele was most strongly linked with rs2242652-A (r^2^=0.62) and rs10069690-T (r^2^=0.48, **Table S5, Figure S6**), suggesting that it might contribute to the associations detected for the GWAS signals for rs2242652 and rs10069690.

### VNTR6-1 creates an expandable G quadruplex that modulates *TERT* splicing

VNTR6-1 is located ∼3.5 kb upstream of *TERT* exon 7 (**Figure 2a**). The simultaneous inclusion or skipping of exons 7 and 8 defines the *TERT* full-length (*TERT-FL)* or *TERT-β* isoform, respectively^17^. To assess the functional effect of VNTR6-1 on *TERT* splicing, we deleted the entire VNTR6-1 region (2,241 bp in the reference human genome) by CRISPR/Cas9 editing.

**Figure 2.**
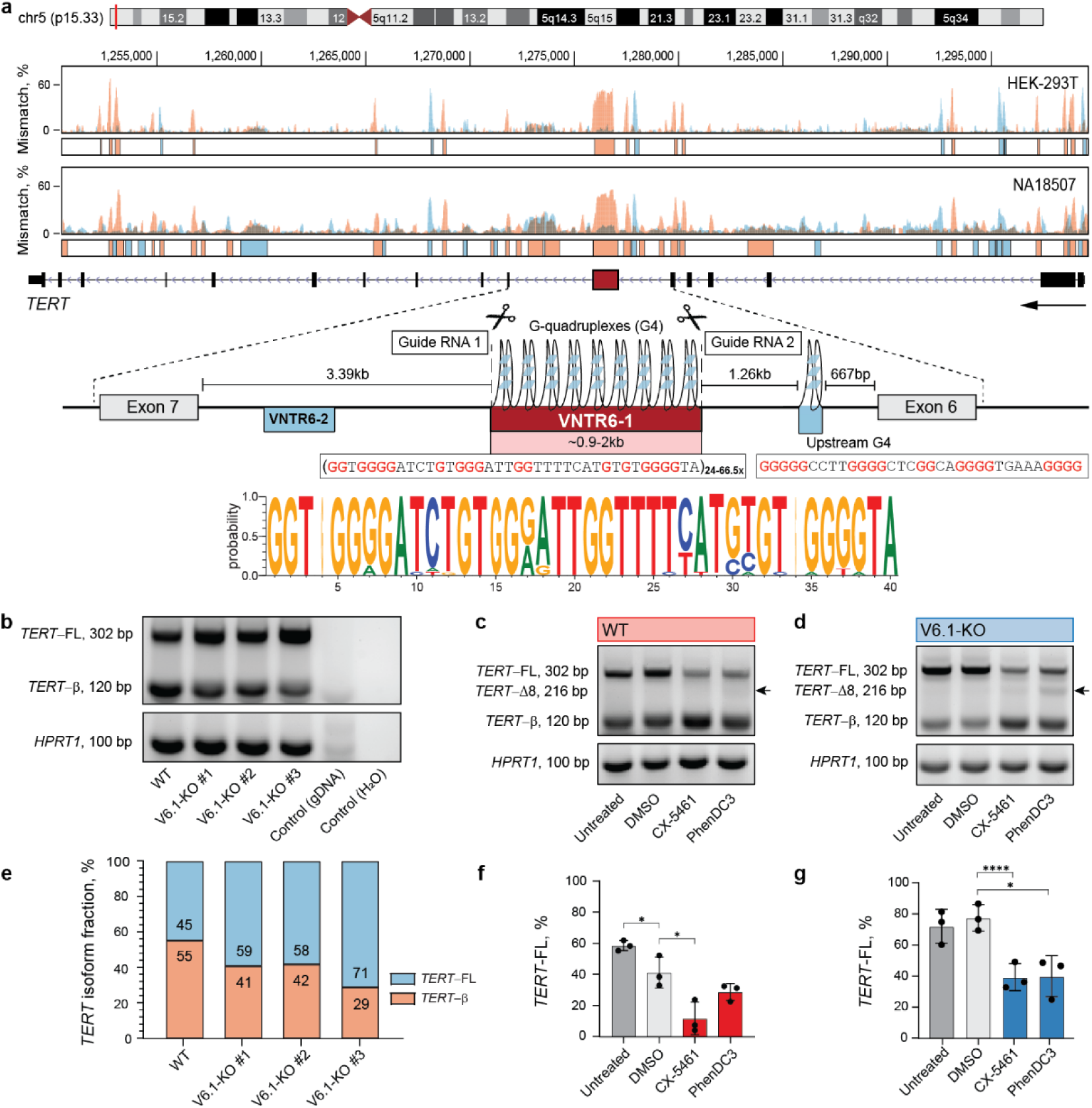
VNTR6-1 affects the splicing ratios of the *TERT-FL* and *TERT-β* isoforms. **a**, G4-ChIP results within the *TERT* region in the HEK-293T (VNTR6-1: Long/Long) and NA18507 (YRI, 1000G, VNTR6-1: Short/Short) cell lines display mismatches (%) during DNA synthesis, reflecting polymerase stalling after G stabilization in both the plus (blue) and minus (orange, direction of *TERT* transcription) genome strands. The genomic region of *TERT* intron 6 shows VNTR6-1 (24-66.5 copies of the 38-bp repeat unit), VNTR6-2, G4s in the minus strand (polymorphic G4 within VNTR6-1 and constitutive upstream G4), and CRISPR/Cas9 guide RNAs for excising VNTR6-1. The sequence logo shows the consensus of the 38-bp VNTR6-1 repeat unit in UMUC3 cells based on PacBio long-read WGS. **b**, Agarose gels of RT‒PCR products amplified from cDNA of corresponding samples; gDNA–genomic DNA was used as a negative control; *HPRT1* was used as a normalization control. **e**, Densitometry results of the PCR amplicons in plot **b**. The differences in the *TERT* isoform ratios are further explored in **Figure S11**. Experiments in UMUC3 cells comparing *TERT* splicing and isoform-specific expression after 72 hrs of treatment with G4 stabilizing ligands, normalized to *HPRT1* as an endogenous control in the WT (**c**, **f**) and V6.1-KO (**d**, **g**) cell lines. **c**, **d**, A representative agarose gel of SYBR-Green RT‒qPCR products detecting several isoforms with primers located in exons 6 and 9. The extra PCR band, marked by an arrow in panels **c** and **d**, is further explored in **Figure S12**. **f**, **g**, Densitometry analysis of the corresponding agarose gels evaluating the *TERT*-*FL* (%) relative to the total PCR products. All analyses are based on three experiments, with one representative gel shown. Comparisons were made against the vehicle control (DMSO). Statistical significance is indicated as follows: **p < 0.01, ***p < 0.001, ****p < 0.0001, Student’s T-test.

Partial deletion of this highly repetitive genomic region was technically impossible. We established three stable isogenic knockout clones (V6.1-KO) in UMUC3, a bladder cancer cell line with high *TERT* expression (DepMap transcripts per million [TPM]=6.78 (**Figure 2a**, **Figure S7a-b**) and two clones in A549, a lung cancer cell line with moderate *TERT* expression (TPM=3.63, **Figure S8a**). Deletion of VNTR6-1 increased the inclusion of exons 7 and 8, shifting the ratio of TERT-FL from ∼45% to 71% in UMUC3 (**Figure 2b, 2e, Figure S7c-f**) and from 34% to 49% in A549 (**Figure S8b-c**). These results suggest that VNTR6-1 acts as a splicing switch between the *TERT-FL* (expressed at a higher fraction in V6.1-KO cells) and *TERT-β* (expressed at a higher fraction in WT cells).

We found no evidence of differential DNA methylation (**Figure S9**) or long-range chromatin interactions (**Figure S10**) involving the VNTR6-1 region. However, we noted a high G content within the 38-bp consensus repeat sequence of VNTR6-1 (5’-GGTGGGGATCTGTGGGATTGGTTTTCATGTGTGGGGTA-3’). Based on G4Hunter analysis and G4-ChiP-seq, we predicted that VNTR6-1 could adopt a G quadruplex (G4) structure in the *TERT*-sense orientation, creating 35-113 G4s per allele with conserved core G-containing motifs (**Figure 2a, Figure S11a, b**).

As a single invariable G4 upstream of VNTR6-1 has been implicated in *TERT-β* splicing^18^, we hypothesized that the variation in the number of G4s, created by VNTR6-1-Short vs. Long alleles, could affect splicing and the *TERT-FL*:*TERT-β* isoform ratio. We treated our isogenic UMUC3 and A549 cell lines—WT, representative of the VNTR6-1-Long allele (**Figure 2c, 2f, Figure S11c-d** for UMUC3**; Figure S8d, e, h, i** for A549) and V6.1-KO, representative of VNTR6-1-Short allele (**Figure 2d, g, Figure S11e-f** for UMUC3; **Figure S8f, g, j, k** for A549), —with several G4-stabilizing ligands. In cDNA from the treated and untreated cells, we quantified the expression of exons 6-9 (*TERT-β*) and 7-8 (*TERT-FL*) and total *TERT*. Treatment with G4 ligands Pidnarulex (CX5461)^19^ or PhenDC3^20^ decreased the *TERT-FL* fraction while increasing the *TERT-β* fraction in both the WT and V6.1-KO cell lines, likely by stabilizing VNTR6-1-G4s (**Figure 2f, g**). In UMUC3, CX5461 increased the *TERT-β* fraction 4.7-fold in the WT cells compared to 2.9-fold in the V6.1-KO cells **(Figure S11c, S11e**). PhenDC3 also significantly increased total *TERT* expression in WT cells, whereas no changes were observed in V6.1-KO **(Figure S11d, f**). These results suggest that while invariable G4s in intron 6 might influence the baseline level of *TERT* splicing, the G4s formed by VNTR6-1 further modulate these splicing ratios and total *TERT* expression. A novel splicing isoform with exon 8 skipping (*TERT*-Δ8, **Figure S12**) was observed in V6.1-KO and WT cells after ligand treatment.

### rs10069690-T and VNTR6-1-Long alleles affect *TERT* expression and splicing

*TERT* expression is generally low in normal human tissues (The Genotype-Tissue Expression (GTEx) Project, median TPM=0.00-2.73) and is not associated with the GWAS leads rs2242652 and rs10069690 (**Table S7**). However, *TERT* expression is generally higher in tumors (The Cancer Genome Atlas (TCGA), median TPM=0.02-5.71; **Table S7**) and is associated with these SNPs in some tumor types (kidney chromophobe, KICH and head and neck squamous carcinoma, HNSC; **Table S7**). We detected high *TERT* expression (mean TPM=59.7, **Figure 3a, Table S8**) in a set of 78 Burkitt lymphoma (BL) tumors^21^. BL is an aggressive pediatric cancer originating from germinal center B cells, in which high TERT expression is necessary for the longevity of memory B cells^22^. Two hotspot somatic mutations in the *TERT* promoter, C228T (− 124 bp) and C250T (−146 bp), upregulate *TERT* expression in many tumors^23,24^, but these mutations are absent in non-Hodgkin lymphomas, including BL^25^ and our set of BL tumors^25^. The combination of high *TERT* expression in the absence of upregulating promoter mutations in BL tumors provides an opportunity to explore the regulation of *TERT* expression by germline variants.

**Figure 3.**
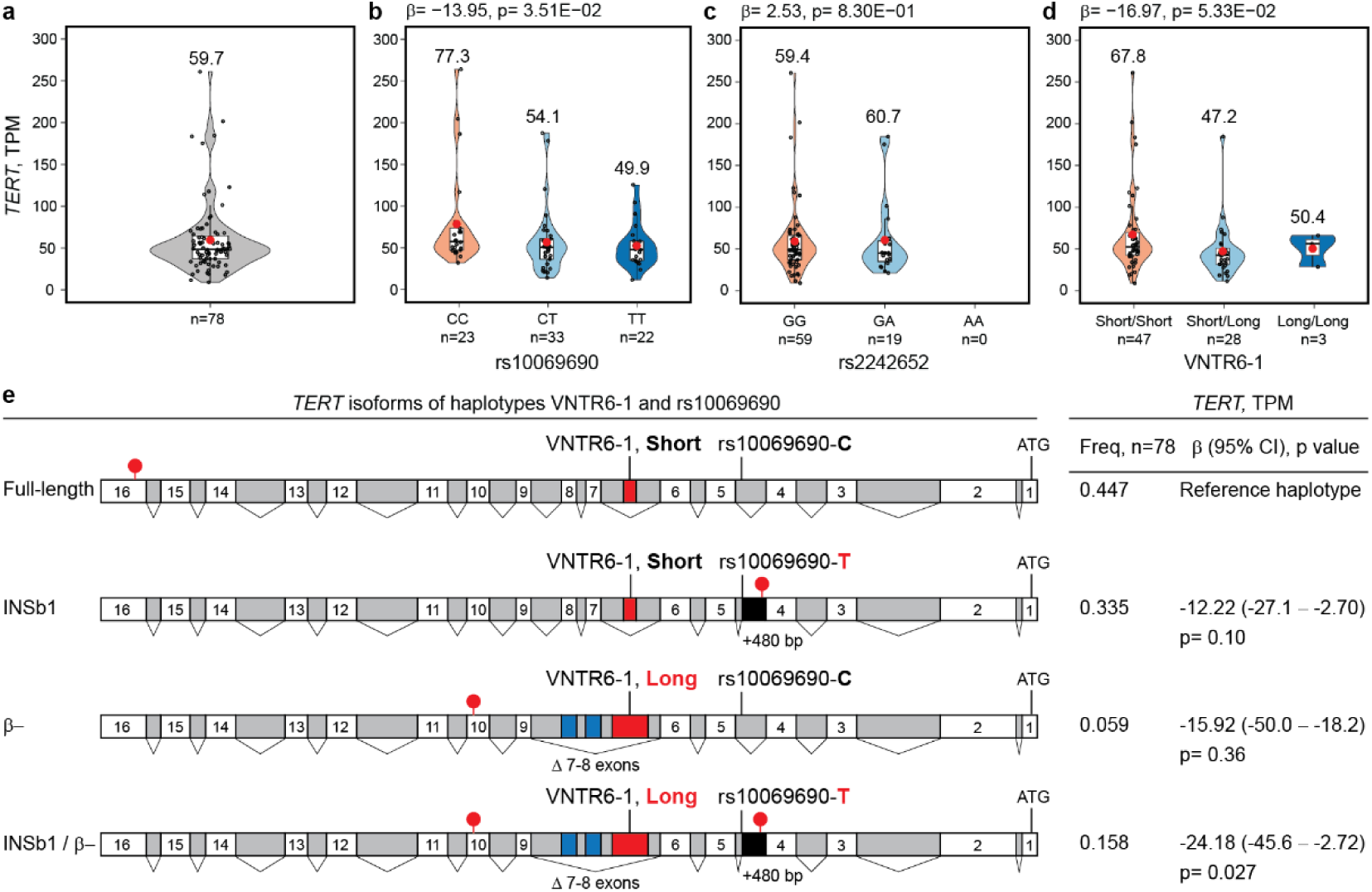
Analysis of *TERT* expression in 78 Burkitt lymphoma (BL) tumors. Total *TERT* expression analyzed as transcripts per million (TPMs) **a**, overall and in relation to the **b**, rs10069690, **c**, rs2242652 and **d**, VNTR6-1 genotypes. Group means are shown as red dots with values above corresponding violin plots. **e**, association of the VNTR6-1 and rs10069690 haplotypes with total *TERT* expression. The reference Short-C haplotype corresponds to the telomerase-encoding *TERT-FL* isoform, while the *INS1* and *TERT-β* isoforms encode truncated proteins without telomerase activity. Effect alleles in haplotypes are marked in red; white boxes – exons; gray boxes – introns; black boxes – intron 4 retention; blue boxes – alternative exons 7 and 8; and red lollipops – stop codons. The direction of the *TERT* exons is from right to left, corresponding to the minus strand, as presented in the UCSC browser. “ATG” indicates translation start codons. P values and β-values are for linear regression models adjusted for sex and age.

In BL tumors (**Table S8**), *TERT* expression decreased with the rs10069690-T allele (β=−13.95, p=0.035; **Figure 3b**) but not with the rs2242652-A allele (β=2.53, p=0.83; **Figure 3c**), with a suggestive trend for decreased *TERT* expression associated with the VNTR6-1-Long allele (β=−16.97, p=0.053; **Figure 3d**). These variants are in high LD in 1000G-EUR but in low LD in 1000G-AFR and our set of BL tumors (88% from African patients, **Figure S13**), suggesting independent effects of rs10069690 and VNTR6-1 on *TERT* expression. Based on the LD profiles and association with *TERT* expression in BL tumors, we functionally prioritized rs10069690 and VNTR6-1 for further analyses.

The functional role of the rs10069690-T allele has previously been attributed to the creation of an alternative splicing site in the *TERT* intron 4, resulting in the coproduction of telomerase-functional TERT-FL and a truncated telomerase-nonfunctional INS1b isoform^26^. However, due to the low *TERT* expression in most human tissues, this relationship has not been explored in relation to genetic variants in the 5p15.33 region^26^. In BL tumors, 26.2% of all RNA-seq reads between exons 4 and 5 were retained within intron 4, in contrast with neighboring introns 3 and 5 (with 10.5% and 8.1% of the retained reads, respectively) (**Figure S14a**). The rate of *TERT* intron 4 retention was more statistically significantly associated with rs10069690 (p=5.36E-09, **Figure S14b**) than with rs2242652 (p=5.0E-03, **Figure S14c**). We analyzed four splicing events between exons 4 and 5, one with canonical intron 4 splicing and three with intron 4 retention (isoforms *INS1*^17,26^, *INS1b*^26^ and with unspliced intron 4, **Figure S15a-d**). The fraction of canonical intron 4 splicing decreased (68.3%, 63.8% and 57.3% of the reads in the rs10069690-CC, CT and TT genotype groups, respectively; p=1.65E-05; **Figure S15e**). These results were consistent with previous observations on the association of the rs10069690-T allele with *INS1b*-type splicing^26^; in BL tumors, the fraction of *INS1b* splicing increased (0% to 3.8% and 7.0% of all reads between exons 4 and 5 in the rs10069690-CC, CT and TT genotype groups, respectively; p=3.07E-09; **Figure S15e**). The fraction of *INS1* was decreased, and that of unspliced intron 4 (excluding reads for *INS1* and *INS1b* isoforms) was increased with the rs10069690-T allele (**Figure S15h**). These results suggest that *INS1-* and *INS1b-*type splicing is minor and could be secondary to intron retention, which increases with the rs10069690-T allele.

Several other common *TERT* isoforms have been reported^17^ (**Figure S16**). The *TERT-α* isoform involves in-frame 36-bp skipping within exon 6 (Δ6_(1–36)_), causing partial loss of the reverse transcriptase domain^17^. As discussed above, *TERT-β* (Δ7–8)^17^ results from the simultaneous skipping of exons 7 and 8 (182 bp), terminating the frameshifted protein in exon 10.

Additionally, *TERT-αβ* results from concurrent Δ6_(1–36)_ and Δ7-8 splicing events. The expression of these *TERT* isoforms was not significantly associated with rs10069690, rs2242652, or VNTR6-1 in BL tumors (**Table S8**). Transcripts truncated by premature termination codons (**Figure S16**), including *INS* (truncated within exon 5), *INS1b* (intron 4), and *TERT-β* or *TERT-αβ* (exon 10), are likely to be eliminated by nonsense-mediated decay (NMD), reducing total *TERT* expression. Escaping NMD would result in alternative TERT proteins without telomerase activity but still binding the telomerase RNA component (hTR), thus producing dominant-negative competitors of the telomerase-functional TERT-FL.

Due to premature termination codons (in intron 4 for *INS1b* and in exon 10 for *TERT-β*), both rs10069690 and VNTR6-1 increase the fraction of NMD-targeted transcripts encoding telomerase-nonfunctional proteins, decreasing total *TERT* expression and the fraction of the telomerase-encoding *TERT-FL* isoform. To assess the combined effects of these variants, we analyzed *TERT* expression based on the VNTR6-1/rs10069690 haplotypes (**Figure 3e**).

Compared to the Short-C haplotype, the *TERT* expression was decreased by the Short-T (β=−12.2, p=0.10) and Long-C (β=−15.92, p=0.36) haplotypes, with a greater decrease occurring when both the VNTR6-1-Long and rs10069690-T alleles were included in the same haplotype (Long-T, β=−24.18, p=0.027, **Figure 3e, Table S8**). Thus, the decrease in total *TERT* expression and increase in alternative isoforms through different splicing events are independently contributed by both alleles, VNTR6-1-Long and rs10069690-T, with a stronger effect when these events are combined.

### VNTR6-1 contributes to proliferative and anti-apoptotic responses to external stimuli

Differences in the activities and ratios of TERT-FL to TERT-β isoforms could contribute to variability in intracellular dynamics and alter cell phenotypes. We compared cell proliferation rates of WT and V6.1-KO UMUC3 cell lines by two methods: a real-time cellular impedance system and a flow cytometry assay based on the dilution of an intracellular dye. To mimic some potentially relevant environmental exposures, cell lines were cultured in media supplemented with either full fetal bovine serum (full serum) or charcoal-stripped serum (CS serum, depleted of hormones and growth factors), (**Figure 4a-c**). Under both serum conditions, the V6.1-KO cells grew slower than WT cells. While both cell lines grew faster in full serum than in CS serum, the increase in proliferation stimulated by full serum was significantly lower in V6.1-KO than that of WT cells (**Figure 4a, Table S9**). Proliferation rate differences between WT and V6.1-KO cells were also apparent after a short period (24 hours) of serum starvation prior to seeding (**Figure S17c-d**), further suggesting that VNTR6-1 functions are sensitive to environmental signals in culture media. Continuous culturing resulted in an increased *TERT-β* fraction (**Figure S17e-h**), potentially reducing proliferation due to limited nutrients as the cells reached confluency. Total *TERT* expression was not affected by the increase in cell density in WT cells but decreased in V6.1-KO cells (**Figure S17f, h)**. This finding suggested that VNTR6-1 might modulate cellular proliferation in response to environmental signals, including hormones and growth factors, and cell density.

**Figure 4.**
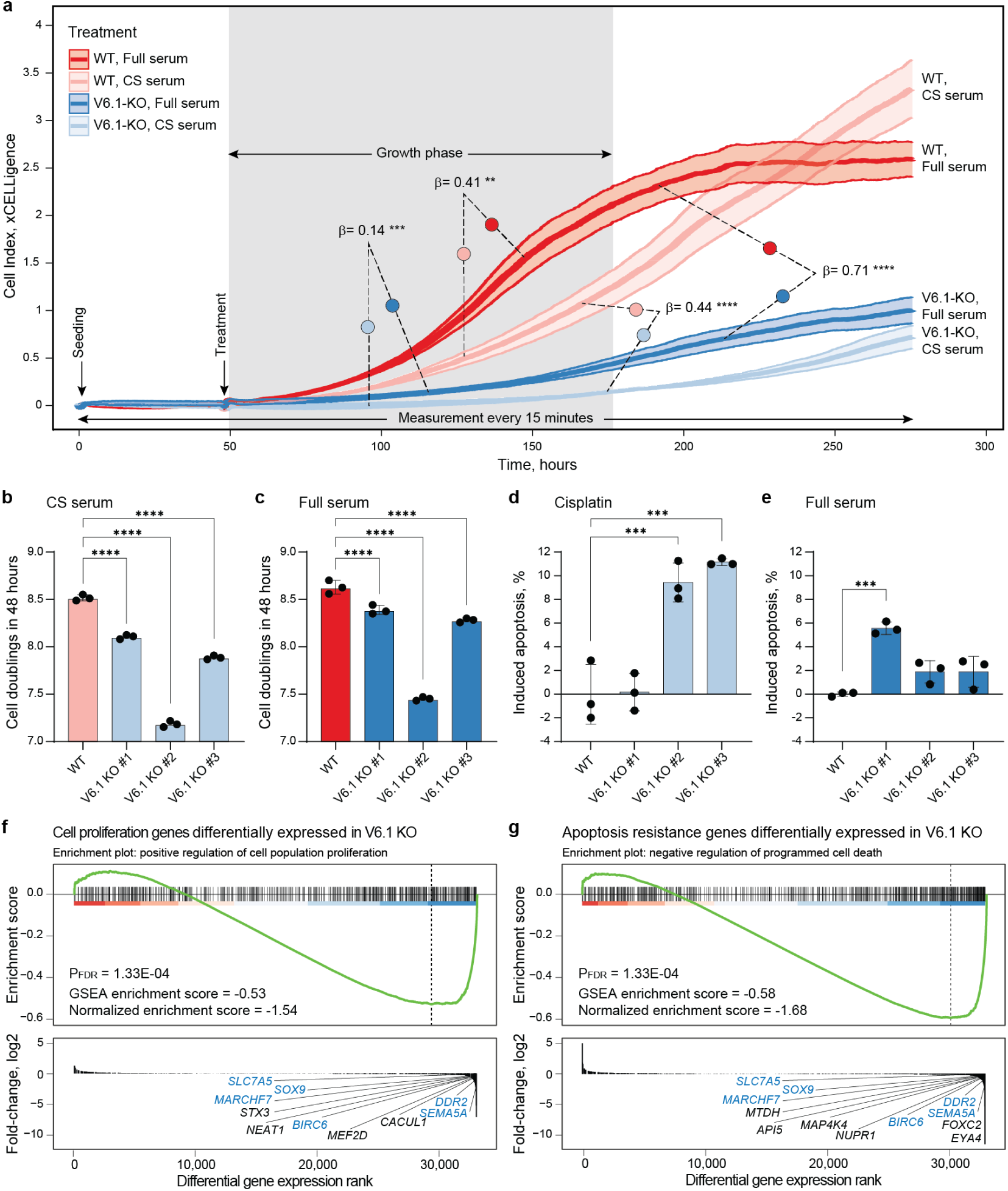
VNTR6-1 affects the proliferation and apoptosis of UMUC3 cells. **a**, Real-time monitoring of cell population growth dynamics (cell index) for 283 hrs in UMUC3 WT and V6.1-KO cells cultured in media supplemented with full serum or charcoal-stripped (CS) serum revealed significantly greater proliferation rates in WT cells than in V6.1-KO cells under both culture conditions. Statistical significance and β-values for differences in the cell index during the visually determined growth phase (gray highlighting between 50 and 183 hours) were calculated using linear mixed-effects models based on six replicates, **p < 0.01, ***p < 0.001, ****p < 0.0001. **b**, Quantification of cell doubling events in CFSE-stained cells cultured with CS serum or **c**, full serum medium for four days in three replicates (****p < 0.0001, Student’s t test). **d**, Cells were treated with 10 µM cisplatin or **e**, full serum medium for 48 hrs, followed by Annexin V-FITC staining to determine the percentage of apoptotic cells in three replicates (***p < 0.001, Student’s t test). Differential expression of genes involved in pathways related to the downregulation of **f**, cell proliferation (positive regulation of cell population proliferation pathway, GO:0008284, **Table S11**) and **g**, apoptosis resistance (negative regulation of programmed cell death pathway, GO:0043069, **Table S11**) according to RNA-seq analysis of V6.1-KO UMUC3 cells compared to WT UMUC3 cells. Genes highlighted in blue are common to both pathways. The data shown in all panels except **f** and **g** represent one of three independent experiments.

Additionally, flow-based apoptosis analyses demonstrated a significant increase in the percentage of apoptotic cells in V6.1-KOs compared to WT cells. This was evident when cells were in full serum and stimulated to grow or upon cisplatin treatment, which induces cell death (**Figure 4d, 4e**). RNA-seq analysis also demonstrated enrichment in both apoptosis and proliferation pathways in V6.1-KO compared to WT cells (**Figure 4f, Table S10, S11**).

TERT-β is a dominant-negative competitor of TERT-FL for telomerase function^27^, but the contributions of these isoforms to cell proliferation are less clear. We monitored cell proliferation after transient overexpression of the *TERT-FL* and *TERT-β* isoforms in 5637 cells, a bladder cancer cell line with low *TERT* expression (DepMap TPM=1.23). Overexpression of both isoforms similarly increased cell proliferation compared to that of the GFP control (**Figure S18, Table S12**). In the co-transfection experiments, cell proliferation increased the most at a 50:50% *TERT-FL*:*TERT-β* ratio, followed by at a 20:80% ratio (**Figure S18, Table S12**). The 50:50% ratio appears to be optimal, potentially because it offers a balance between promoting proliferation and reducing cell death.

Structured illumination microscopy fluorescence imaging of A549 cells (a cell line with a large cytoplasm, facilitating visualization) co-transfected with both TERT isoforms revealed stronger mitochondrial co-localization for TERT-β than for TERT-FL (**Figure 5a, Figure S19**). These results further suggest the role of TERT-β in mitochondrial-localized processes, one of which may be protection from apoptosis. The effects on proliferation and apoptosis achieved by VNTR6-1 knock-out (**Figure 5b)** suggest that the ratio of TERT-FL to TERT-β may affect both cellular longevity (by protecting cells from apoptosis) and replicative potential (by altering proliferation in response to environmental conditions).

**Figure 5.**
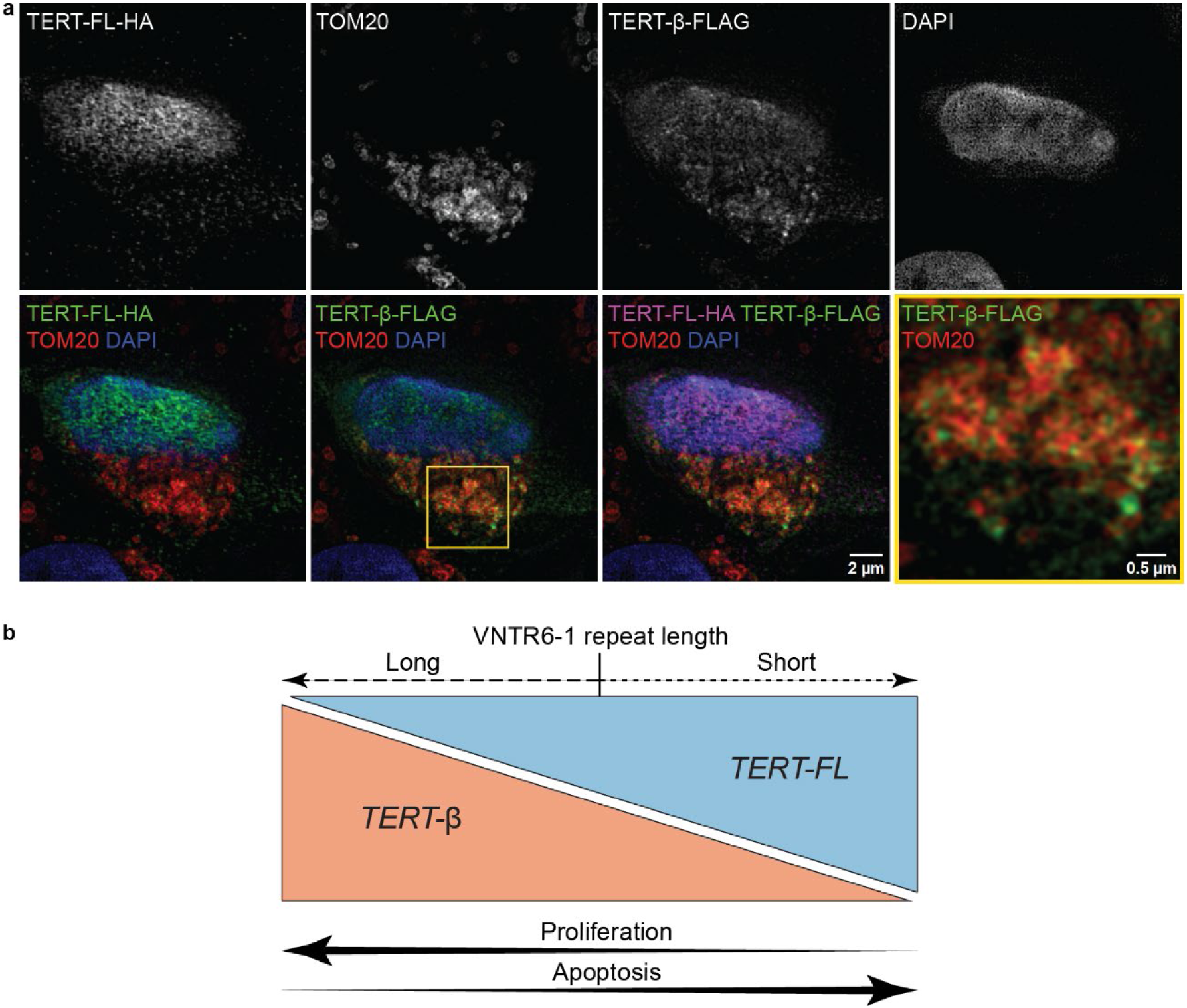
The functional differences between the TERT-FL and TERT-β isoforms. **a,** Structured illumination microscopy images of A549 cells co-transfected with TERT-FL and TERT-β expression constructs at a 50:50% ratio. For individual channels, staining is shown as black/white images for better contrast. On tri-color merged panels, green – FLAG (TERT-β) or HA (TERT-FL), blue – DAPI (nuclei). On the quad-color merged panel, purple - HA (TERT-FL), green - FLAG (TERT-β), red - TOM20 (mitochondria), blue - DAPI (nuclei). The yellow inset in the TERT-β-FLAG panel is shown at a higher magnification to demonstrate colocalization with mitochondria (yellow staining). **b,** The overview of the VNTR6-1, *TERT*-FL:*TERT*-β ratio, proliferation, and apoptosis.

### VNTR6-1, rs10069690 or their haplotypes account for pleiotropic cancer GWAS associations

Because we established that VNTR6-1 is linked with GWAS leads rs2242652-A (r^2^=0.62) and rs10069690-T (r^2^=0.48) in the 1000G-EUR populations, we next sought to compare the associations of these markers with cancer risk. Having validated the rs56345976/rs33961405 haplotypes as a confident predictor of VNTR6-1 Short vs Long alleles (**Table S3, S4, Figure S3, S4**), we used these haplotypes to infer VNTR6-1 alleles in various sets. Specifically, we inferred VNTR6-1 and the composite marker VNTR6-1/rs10069690 because it captured functional effects from both variants. Using these markers, we performed association analyses in individuals of European ancestry from the Prostate, Lung, Colorectal and Ovarian (PLCO) cohort of cancer-free controls (n=73,085) and 29,623 patients with 16 cancer types^28^. The PLCO association results for the VNTR6-1-Long allele and VNTR6-1/rs10069690 were comparable to those for the rs10069690-T and rs2242652-A alleles; these alleles were associated with a reduced risk of bladder and prostate cancer but an elevated risk of breast, endometrial, ovarian, and thyroid cancer and glioma (**Figure 6a, Table S13**). Only for prostate cancer the association was stronger for rs10069690 than for VNTR6-1 and rs22942652, perhaps suggesting a more important role of decreased *TERT* expression due to intron 4 retention and *INS1b*-splicing in the molecular mechanism of this cancer. Compared to the reference Short-C haplotype, the strongest positive or negative cancer-specific associations were for the Long-T haplotype (**Figure S20**). The SNPs capturing VNTR6-1 status (rs56345976 and rs33961405) were moderately associated with some cancer types (**Table S13**).

**Figure 6.**
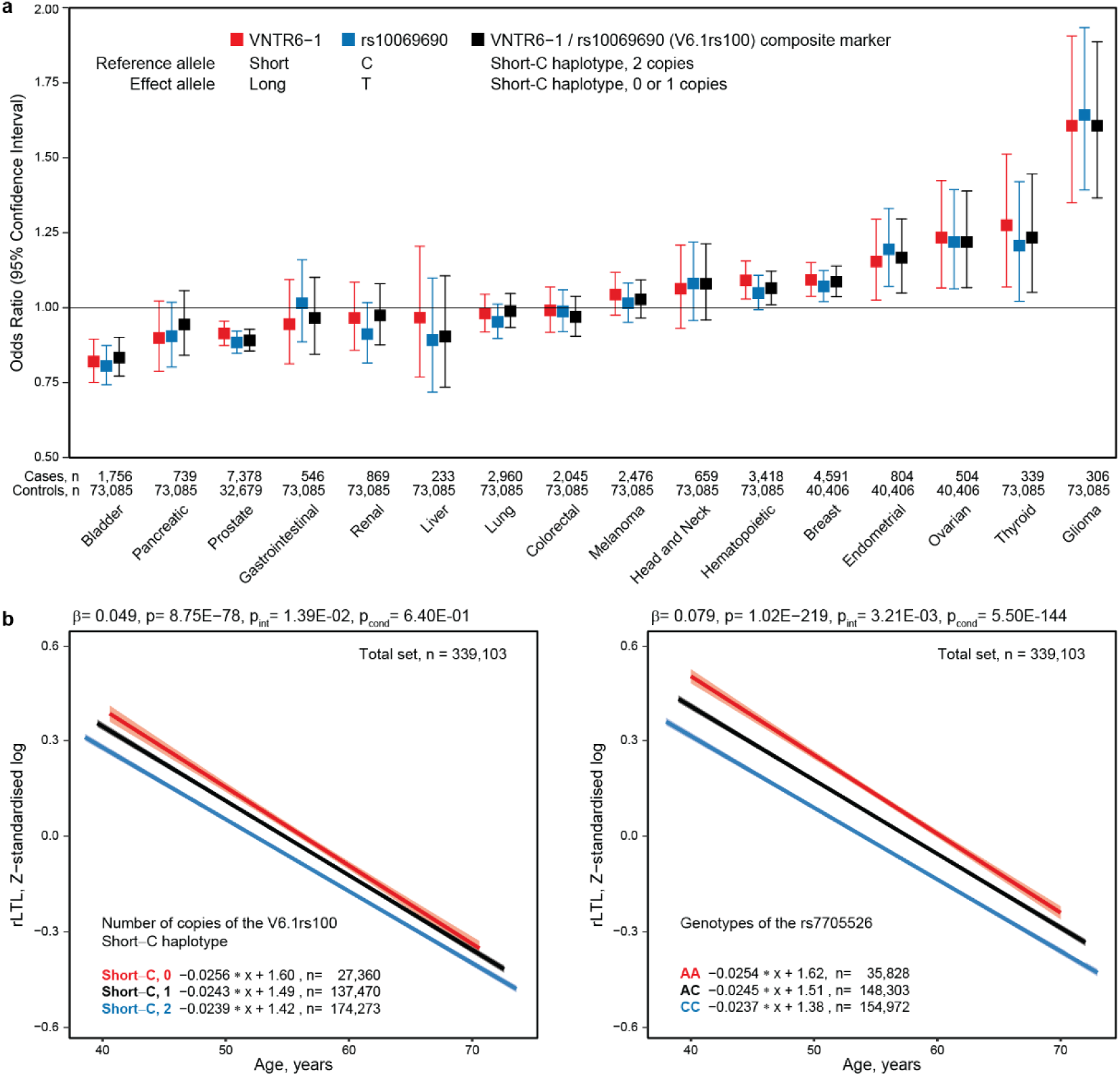
Association analyses for cancer risk in PLCO and relative leukocyte telomere length (rLTL) in UKB cancer-free individuals. **a**, Evaluation of cancer risk associated with the VNTR6-1-Long and rs10069690-T alleles and the composite marker (VNTR6-1/rs10069690) in the PLCO dataset (n=102,708). Odds ratios (ORs) with 95% confidence intervals (CIs) were calculated for comparisons between patients with the indicated cancers and a common group of cancer-free controls using logistic regression analysis with an additive genetic model adjusted for sex and age. **b**, Evaluation of the relationships between rLTL and VNTR6-1/rs10069690 and rs7705526 in UKB cancer-free individuals (n=339,103, LD, r^2^=0.33 between VNTR6-1/rs10069690 and rs7705526). P values and β-values were derived from linear regression models adjusted for sex, age, and smoking status. P_int_ represents the interaction between genotypes and 5-year age groups; P_cond_ represents the mutual adjustment for rs7705526 or VNTR6-1/rs10069690. The graphs display regression lines with 95% confidence intervals and regression equations. The analysis revealed a decrease in the rLTLs with more copies of the Short-C haplotype. The results of sex-specific analyses of VNTR6-1/rs10069690 are presented in **Figure S20b**.

### Associations of *TERT* isoforms and genetic variants with telomerase-related metrics

*TERT-β,* which encodes a telomerase-nonfunctional protein, is the major *TERT* isoform in both normal and tumor tissues (**Figure S20**). In 30 normal GTEx tissue types, the *TERT-FL* and *TERT-β* isoforms represented on average 17.7% and 58.5%, respectively, of the total *TERT* expression, while they represented 38.4% and 41.0%, respectively, in 33 tumor types in TCGA (**Table S14**). To further explore the functional differences between TERT-FL and TERT-β, we assessed four telomerase-related metrics: EXpression based Telomerase ENzymatic activity Detection (EXTEND)^29^, stemness (mRNAsi)^30^, the telomerase signature score^31^ and telomere length in primary tumors^31^ (**Figure S21, Table S15**). In GTEx, significant correlations with the EXTEND signature (positive for *TERT-FL* and negative for *TERT-β*) were observed in four tissues (blood, colon, esophagus and testis). Similarly, in TCGA, most tumors with significant correlations across all metrics showed positive values for *TERT-FL* and negative values for *TERT-β.* In TCGA, of the four metrics, telomere length in tumors showed the weakest correlations with *TERT* isoform expression (**Figure S21**), potentially due to somatic events, including *TERT*-upregulating promoter mutations^23,24^.

We next tested whether VNTR6.1, rs10069690 or their haplotypes are associated with relative leukocyte telomere length (rLTL). We inferred VNTR6-1 and VNTR6-1/rs10069690 as described above in cancer-free individuals of European ancestry (n=339,103) from the UK Biobank (UKB)^32^; the Short-C haplotype was associated with shorter rLTLs (β=−0.049, p=8.75E-78, **Figure 6b, Figure S20b, Table S16**). Significant associations were also observed with several known markers^33,34^ within *TERT* intron 2, including rs7705526 (β=−0.079, p=1.02E-219; **Table S16**); adjustment for rs7705526 eliminated the association of the rLTL with VNTR6-1/rs10069690 (p=0.64). Notably, regression slopes for these markers differed by genotypes (**Figure 6b**). Interaction analysis in 5-year interval groups revealed a significantly slower decrease in the rLTL in younger individuals and a faster decrease in older individuals without the Short-C haplotype (a greater fraction of telomerase-nonfunctional TERT) than in those with this haplotype (p_int_=1.39E-02, **Table S16**). This effect remained unchanged after adjustment for rs7705526 (p_int_=1.38E-02, **Table S16**), which had its own significant interaction (p_int_=3.21E-03, **Table S16**). The rLTL association pattern was consistent in a smaller set of healthy individuals with lymphocyte telomere length measured by flow FISH,^35^ but interaction analysis was limited by sample size and age range (**Table S17**).

### The VNTR6-1-Short and rs10069690-C alleles are human-specific variants

The VNTR6-1 genomic region is absent in non-primate species (**Figure S22**). In primates, the VNTR6-1 consensus repeat sequences in chimpanzee and bonobo are nearly identical to those in humans and more divergent in orangutan, with all these primates carrying the Long-T haplotype (**Figure S22**). In the genomes of archaic humans (Neandertal and Denisova), only Long-T haplotypes were observed (**Figure S23**). Thus, the VNTR6-1-Short and rs10069690-C alleles, as well as the Short-C haplotype that increase the fraction of the telomerase-functional TERT-FL isoform, are human-specific and major or common in all modern human populations (**Table S18**). In cancer-free controls of European ancestry, the Short-C haplotype frequencies were comparable (71.36-72.07%) across 40-to 80-year-old age groups in the UKB and PLCO but decreased to 67% in individuals aged 98-108 years (**Figure S24**). Decreased frequencies of both the rs10069690-C and the VNTR6-1-Short alleles contributed to this difference between centenarians and 40-to 80-year-olds.

## DISCUSSION

Cancer risk is influenced by complex interactions between genetic and environmental factors and further depends on the replicative potential of stem cells^36–38^. We showed that reduced or elevated cancer risk associated with a multi-cancer GWAS locus at chr5p15.33 marked by the SNPs rs2242652 and rs10069690 is related to the genetic regulation of *TERT* splicing by rs10069690 and VNTR6-1, a 38-bp intronic tandem repeat. In all populations, VNTR6-1 status can be confidently inferred as a biallelic marker with a Short allele (24-27 copies) vs. a Long allele (40.5-66.5 copies) based on haplotypes of two common SNPs, rs56345976 and rs33961405. We inferred VNTR6-1 (Short vs Long alleles) alleles and explored their distributions in controls from 1000G populations and in cancer patients and controls of European ancestry.

In Europeans, the GWAS associations for rs2242652-A allele were fully explained by the linked VNTR6-1-Long allele (r^2^=0.62), but the associations for rs10069690-T (r^2^=0.48) were partially explained, suggesting that rs10069690 may have an individual functional effect. While we did not detect any functional properties for rs2242652, we demonstrated that both the VNTR6-1-Long and rs10069690-T alleles independently and their combined haplotype (i.e., the Long-T haplotype) alter the *TERT* isoform ratios by reducing telomere-functional TERT-FL and increasing alternative telomerase-nonfunctional INS1b and TERT-β isoforms. We propose that genetically regulated levels of expression and the ratios of these isoforms affect cellular longevity by protecting cells from apoptosis and altering their replicative potential at homeostasis or in response to environmental factors (**Figure 7**).

**Figure 7.**
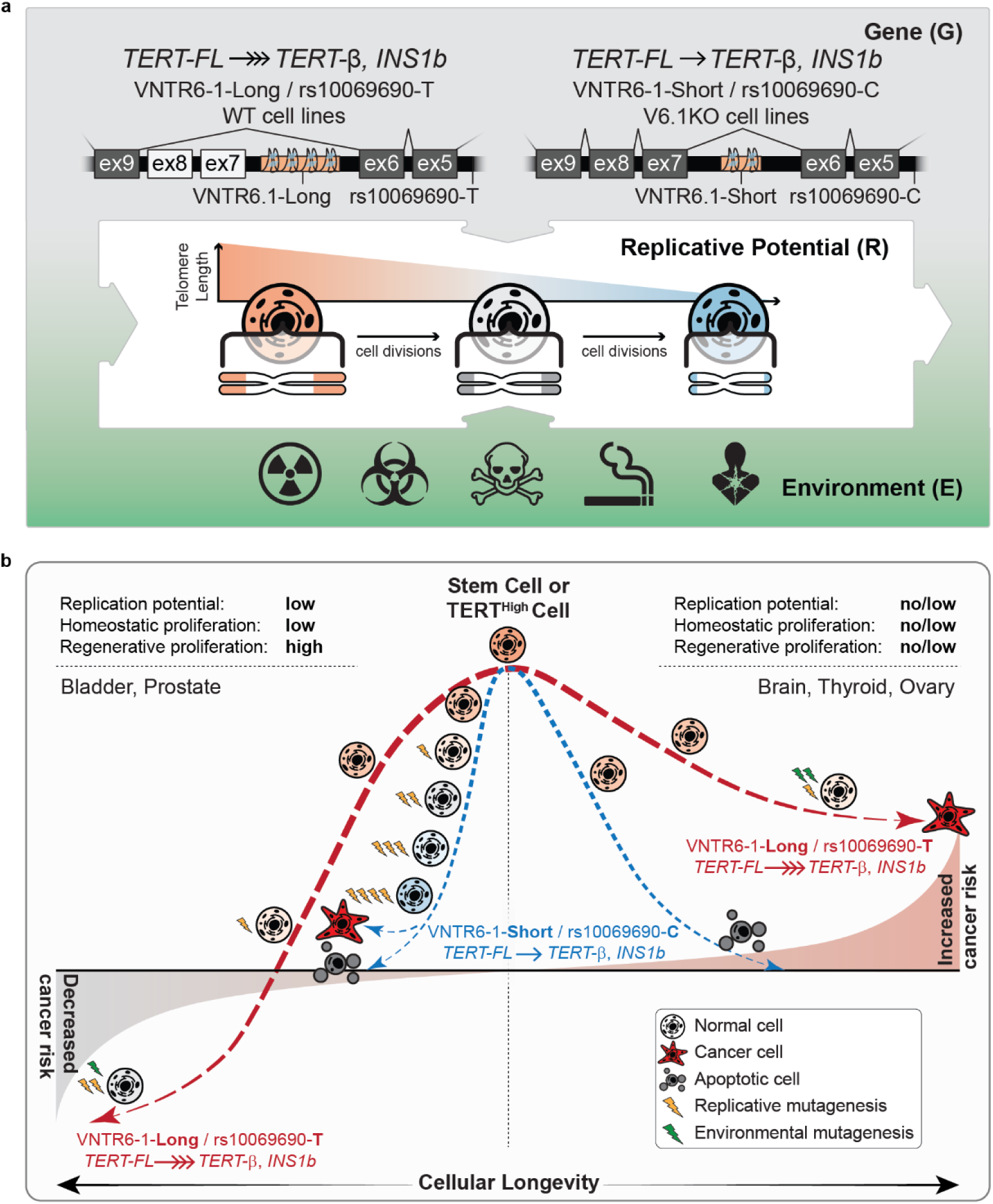
The interaction model of factors affecting cancer risk. **a**, *TERT* genetic variants VNTR6-1 and rs10069690 and environmental factors define the relative ratios of the isoforms encoding telomerase-functional TERT-FL and telomerase-nonfunctional TERT-β and INS1b isoforms. These isoforms affect cell proliferation, apoptosis and telomere length, thus modulating cellular longevity and replicative potential, including homeostatic proliferation, which maintains tissue self-renewal, and regenerative proliferation, which responds to environmental factors and tissue damage. **b**, Cancer risk as a product of G x E x R interactions. The VNTR6.1-Long and rs10069690-T alleles, or their haplotype (Long-T), are associated with reduced cancer risk in tissues with low homeostatic but high regenerative potential (e.g., bladder). The anti-apoptotic effect of the *TERT-β* isoform reduces the need for regenerative proliferation, thus decreasing the risk of acquiring mutations due to replicative mutagenesis. In tissues with no/low homeostatic and regenerative proliferation (e.g., brain, thyroid, ovary), the same alleles and Long-T haplotype are associated with elevated cancer risk. The anti-apoptotic effect of *TERT-β* extends cellular longevity, allowing the accumulation of more mutations due to environmental mutagenesis, such as through exposure to reactive oxygen species (ROS), cellular metabolites, etc.

The alleles/haplotypes reducing the TERT-FL and increasing the TERT-β isoform were associated with an elevated risk of cancers originating from tissues (e.g., brain, thyroid, ovary) with no/very low replicative potential at homeostasis and no regenerative replication, i.e. cell growth to repair the tissue damage. Notably, the *TERT-β* isoform accounts for ∼80% of the total *TERT* expression in these normal tissues (**Table S14**). The anti-apoptotic effect of the TERT-β isoform may extend the lifespan of these cells, allowing for the accumulation of somatic mutations over time, increasing cancer risk.

However, the same alleles/haplotypes were associated with a reduced risk of cancers originating from tissues with low homeostatic proliferation but potentially high regenerative proliferation to repair tissue damage caused by environmental exposures and stressors, including pathogens, hormones and reactive metabolites (e.g., bladder and prostate cancer). In these cases, the anti-apoptotic effect of the TERT-β isoform may reduce the extent of regenerative proliferation, thus limiting mutagenesis caused by replication errors.

The reduced fraction of the telomerase-encoding TERT-FL isoform leads to a decreased availability of the TERT-TERC complex for telomere maintenance, making it less likely for mutated cells to achieve immortalization and expansion. Tumorigenesis in tissues with low replicative potential requires driver mutations, such as *TERT*-upregulating promoter mutations^39^, which can be acquired through replicative or environmental mutagenesis. Rare cells with high TERT expression (TERT-high cells) can serve as stem cells to support tissue regeneration^40^ and initiate tumorigenesis after acquiring driver mutations.

We did not detect GWAS associations for the same alleles/haplotypes for cancers originating from tissues with high homeostatic proliferation (e.g., the gastrointestinal tract). High proliferation rates in stem cells of these tissues, combined with cell death induced by critical telomere shortening in differentiated cells, prevent cells from reaching a malignant state and thus act as a tumor-suppressive mechanism^41^. For cancer types with no or marginal associations for these alleles/haplotypes, TERT-related mechanisms might be more heterogeneous and dependent on cell specificity, tumor subtype, and timing, as well as the type and intensity of environmental exposure.

While the telomerase activity of TERT is essential for cellular homeostasis, non-canonical roles have also been recognized, including roles in the DNA damage response and radiosensitivity^44^. TERT protects mitochondrial function and reduces the production of reactive oxygen species (ROS)^42^. Telomere shortening is accelerated under oxidative stress conditions^42^, potentially through increased damage to telomere DNA or/and the relocation of TERT from the nucleus to mitochondria^43^. Our data and previous observations^27^ of the predominant localization of telomerase-nonfunctional TERT-β to mitochondria support the role of this isoform in cancer risk or protection through telomerase-independent regulation of apoptosis.

Telomere length has been extensively studied in relation to cancer and non-cancer conditions^45,46^. Mendelian randomization analysis revealed an association between genetically predicted longer telomeres and the risk of 8 of 22 cancer types tested, especially for rare cancers and cancers of tissues with low replicative potential^47^. Our analysis in the UKB showed a strong association for the VNTR6-1/rs10069690 haplotypes with rLTL but weaker than for the other *TERT* rLTL markers (rs7705526, rs2736100, and rs2853677) used for predicting telomere length^33,34^. We noted a greater degree of telomere shortening in older than in younger individuals without the Short-C haplotype. This might be due to a greater proportion of circulating lymphocytes originating from stem cells and their progenitors that have experienced more cell divisions due to increased cellular longevity provided by the anti-apoptotic TERT-β isoform.

The alleles associated with an increased ratio of telomerase-encoding *TERT-FL* isoform, VNTR6-1-Short, rs10069690-C, and their Short-C haplotype are human-specific variants with a high frequency in 40-to 80-year-old European individuals but a lower frequency in centenarians. The emergence and retention of these alleles might be consistent with the disposable soma theory of ageing, which postulates that evolution favors factors supporting reproductive fitness and growth at the expense of longevity, which requires substantial maintenance to repair the somatic damage that accumulates with increasing age^48^. Female fertility strongly depends on ovarian telomerase^49^, and telomere shortening is considered an evolutionary cost of reproductive trade-offs^50^. The evolutionary selection of genetic variants that increase the ratio of the telomerase-encoding *TERT-FL* isoform might provide this reproductive fitness benefit while decreasing longevity later in life, perhaps due to elevated cancer risk.

Some limitations of our study include the lack of longitudinal rLTL data and not considering other possible tissue-specific effects of *TERT* splicing regulation by VNTR6-1 and rs10069690. Further studies are warranted to explore our findings in the context of other cancer GWAS signals within the 5p15.33 region^3,4^. In conclusion, we demonstrate that the multi-cancer GWAS locus at 5p15.33 marked by rs10069690 and rs2242652 can be genetically and functionally accounted for by a combination of the SNP rs10069690 within intron 4 and a VNTR within intron 6 (VNTR6-1) of *TERT*. These variants independently regulate *TERT* splicing and expression, with a stronger combined effect. The *TERT* isoform ratios affect cellular longevity and replicative potential, which can be further modulated by environmental factors, thus contributing to reduced or elevated cancer risk.

## Acknowledgments

This work was supported by the Intramural Research Programs of the Division of Cancer Epidemiology and Genetics (DCEG) and the Center for Cancer Research (CCR), National Cancer Institute, and the Center for Alzheimer’s and Related Dementias (CARD) within the Intramural Research Program of the National Institute on Aging and the National Institute of Neurological Disorders and Stroke (1ZIAAG000538). BLGSP was funded in part by the Foundation for Burkitt Lymphoma Research (http://www.foundationforburkittlymphoma.org) and with U.S. Federal funds from the National Cancer Institute, National Institutes of Health, under Contract No. HHSN261200800001E and Contracts No. HHSN261201100063C and No. HHSN261201100007I (DCEG). The presented results are in part based upon data generated by the TCGA Research Network. The work was conducted using the UK Biobank resource (application 92005). The UK Biobank was established by the Wellcome Trust, the Medical Research Council, the United Kingdom Department of Health, and the Scottish Government. The UK Biobank has also received funding from the Welsh Assembly Government, the British Heart Foundation, and Diabetes UK. The CIBMTR is supported primarily by the Public Health Service U24CA076518 from the NCI, the National Heart, Lung and Blood Institute (NHLBI), and the National Institute of Allergy and Infectious Diseases (NIAID); 75R60222C00011 from the Health Resources and Services Administration (HRSA); and N00014-23-1-2057 and N00014-24-1-2057 from the Office of Naval Research. The Cancer Genomics Research (CGR) Laboratory and Genome Modification Core are funded with Federal funds from the National Cancer Institute under Contract No. 75N910D00024. BP and MM acknowledge the support of the Chan Zuckerberg Initiative and the National Institutes of Health grants U24HG011853 and OT2OD033761 to BP. Max Hogshead was supported by the NCI Intramural Continuing Umbrella for Research Experiences (iCURE) program. We thank Drs. Helen Piontkivska, and the members of the Laboratory of Translational Genomics for comments and discussions. We thank Dr. Tatiana Karpova, Optical Microscopy Core (NCI/CCR/LRBGE), for help with super-resolution imaging. The opinions expressed by the authors are their own and should not be interpreted as representing the official viewpoint of the U.S. Department of Health and Human Services, the National Institutes of Health or the National Cancer Institute.

## Author contributions

O. F-V and LP-O conceived the study; O. F-V, C-H. L and CZ performed the data analysis; MH, MH, BWP and KF performed the experiments; CB, KJB, MK, MM and BP generated the long-read genome assemblies; KF, MH, KJ, WL and KT performed the targeted PacBio sequencing; RC, JS, MJM, SJC, SG, SAS and SMM provided reagents, data, samples and interpretations of the results; O. F-V and LP-O wrote the manuscript with the input of all the authors; and LP-O supervised the project.

## Data availability

All sequencing data generated in this study (PacBio targeted sequencing, PacBio-WGS, HiChIP, RNA-seq) are deposited in the Sequence Read Archive (SRA) under accession numbers (PRJNA1134701 and PRJNA1134698 available at publication). The publicly available datasets used in the study are listed in **Table S19**. The derived data for the public datasets (1000G) are provided in the supplementary tables.

## Code availability

The pipeline and script used for the analysis of genome assemblies are available at GitHub (https://github.com/oflorez/HumanGenomeAssemblies).

Supplementary Information is available for this paper.

Correspondence and requests for materials should be addressed to LP-O.

## ONLINE METHODS

### Human samples used for targeted PacBio-seq and TaqMan genotyping of select SNPs

DNA samples for HapMap I (CEU panel for CEPH Utah residents with ancestry from Northern and Western Europe, n=90), HapMap III (YRI panel for Yoruba in Ibadan, Nigeria, n=90), select samples from the Human Pangenome Reference Consortium (HPRC, n=10), and the European panel of the Georgia Centenarian Collection (n=100) were purchased from the Coriell Institute for Medical Research; deidentified tissue samples for bladder tumors and matching adjacent normal samples (n=5 pairs) were purchased from Asterand Bioscience and used for DNA extraction and genotyping. Flow FISH telomere length samples (n=77) were obtained from donors of hematopoietic cell transplants from the Center for International Blood and Marrow Transplant Research (CIBMTR; https://cibmtr.org) biorepository. Telomere length was measured for total lymphocytes and lymphocyte subsets using the flow FISH assay in a previous study^1^. For the current analysis, the samples were selected to represent a wide range of telomeres (4.5 to 11.2 kb) and telomere length was analyzed in relation to *TERT* genetic variants using linear regression models adjusted for age and sex.

### Cell lines

The urinary bladder cell lines UMUC3 (CRL-1749), 5637 (HTB-9), HT1376 (CRL-1472), RT4 (HTB-2), T24 (HTB-4), and SCaBER (HTB-3), as well as the Burkitt lymphoma cell line Raji (CCL-86) and the lung cancer cell line A549 (CCL-185), were purchased from ATCC (Manassas) and maintained in the recommended media supplemented with 10% FBS (unless specified otherwise) and 1% antibiotics. All cell lines were regularly tested for Mycoplasma contamination using the MycoAlert Mycoplasma Detection Kit (Lonza) and authenticated by Identifiler (Thermo Fisher). For experiments with cells grown in media containing charcoal-stripped (CS) serum, 3-4 days prior to the experiments, the media was changed to phenol red-free EMEM supplemented with 10% CS FBS, 1% GlutaMAX, and 1% antibiotics.

### Analyses of BL tumors

RNA-seq and DNA-WGS data (Illumina) for Burkitt lymphoma (BL) tumors were obtained from the National Cancer Institute (NCI) Cancer Genome Characterization Initiative (CGCI): Burkitt Lymphoma Genome Sequencing Project (BLGSP)^2,3^, dbGaP phs000527.v6.p2. The datasets were accessed through the National Cancer Institute Genomic Data Commons (GDC, https://gdc.cancer.gov/). RNA-seq BAM files were analyzed using read counts based on the R package FeatureCounts (v2.0.6). Splicing events between *TERT* exons 4 and 5 were annotated based on a custom GTF annotation file to perform read summarization at the feature level, generating a raw count matrix. The total number of reads was determined by counting the reads mapped to the splicing junction between exons 4 and 5 and those that extended into intron 4 by at least 20 bp. Read counts were calculated for the splicing events *INS1* (a 38-bp extension of exon 4 into intron 4), *INS1b* (a 480-bp extension of exon 4 into intron 4) and unspliced intron 4 (total reads between exons 4 and 5 minus reads for *INS1* and *INS1b)* as fractions of the read counts for these events within total read counts. BAM files were also used to estimate the overall expression of *TERT* isoforms –α, –β, and –α–β, which were indexed in a GTF file from ENSEMBL and analyzed using MISO (v0.5.4) with default parameters. Transcripts per million (TPM) values for bulk *TERT* RNA-seq data were downloaded from the GDC data portal. eQTL analyses were performed under additive genetic models using the ‘lm’ function in R (v4.3.0), with adjustments for sex and age. *TERT* intron retention was analyzed with IRFinder (v2.0.1) with default settings using the GRCh38 reference genome FASTA file and transcriptome GTF file for annotation.

### Analysis of long-read sequences

VNTR6-1 and VNTR6-2 within *TERT* intron 6 were explored based on long-read sequencing data. Phased genome assemblies for 47 individuals (94 chromosomes) were downloaded in FASTA format from the Human Pangenome Reference Consortium (HPRC)^4^. Additionally, we used 358 long-read sequencing (R9, Oxford Nanopore) DNA assemblies generated by the Center for Alzheimer’s and Related Dementias (CARD) of the National Institute on Aging. Input DNA was extracted from the brain tissue of 179 neurologically normal individuals of European ancestry (<u>dbGaP</u> phs001300.v4.p1) and phased assemblies were generated using the Napu pipeline^5^.

From the assemblies, the genomic sequences were extracted in FASTA format using Cutadapt (v4.0) based on two sets of nested sequences flanking the region of interest, ∼9 kb, GRCh38, chr5:1,271,950-1,281,050. The extracted sequences were aligned to the GRCh38 reference genome using minimap2 (v2.26) and combined in one BAM file, with each individual represented by two sequences, one for each chromosome. In this BAM file, SNPs were scored using SAMtools with mpileup flag (v1.17), and VNTRs were scored using Straglr (v1.4) with default settings. The pipeline is available at https://github.com/oflorez/HumanGenomeAssemblies.

### Targeted PacBio-seq

PCR amplicons for targeted PacBio sequencing of VNTR6-1 were generated using the LA Taq Hot-Start DNA Polymerase Kit (Takara) and M13-tagged primers VNTR6-1-M13F) and VNTR6-1-M13R (**Table S20**). In the reference human genome, these primers capture a genomic fragment of 2,241 bp. The optimized 20 µl reactions included 4% DMSO, 0.3 µl of LA Taq DNA Polymerase, 2.5 µl of 10x LA Taq PCR Buffer, 4 µl of 2.5 mM dNTPs, 0.5 µl of each 10 µM primer, and 25 ng of genomic DNA. The PCR conditions included denaturation for 1 min at 94°C, 36 cycles of denaturation for 10 s at 98°C and combined annealing/extension for 3.5 min at 68°C, followed by a final extension for 10 min at 72°C.

The controls included 1000G DNA samples purchased from the Coriell Institute for Medical Research and selected to represent various repeat lengths determined based on HPRC assemblies (HG00741, HG01358, HG01891, HG02080, HG02622, HG02717, HG02723, HG03453, HG03492, and NA18906) (**Table S2**). For technical validation of the first and second rounds of PCR, all products were quantified with the Quant-iT PicoGreen dsDNA Assay (Invitrogen), and 5% of the products were analyzed with the TapeStation D5000 Kit (Agilent). The second round of PCR was performed with the LA Taq Hot-Start DNA Polymerase Kit and the SMRTbell Barcoded Adapter Complete Prep Kit (PacBio), and the M13 tags incorporated by the first PCR were used to attach unique barcodes to each sample with primers M13F and M13R, where “N” represents the unique barcode (**Table S20**). The 25 µl PCRs included 4% DMSO, 0.4 µl of LA Taq DNA Polymerase, 2.5 µl of 10x LA Taq PCR Buffer II, 4 µl of 2.5 mM dNTPs, 1.0 µl of each 3 µM barcoded M13 primer, and 25 ng of product from the first PCR. The PCR conditions included denaturation for 1 min at 94°C, 10 cycles of denaturation for 10 s at 98°C and combined annealing/extension for 3.5 min at 68°C, followed by a final extension for 10 min at 72°C. The final amplicons from three 96-well PCR plates (288 samples) were pooled, processed with the Sequel II binding kit 3.1 (PacBio), and sequenced on one SMRT Cell on the Sequel II System (PacBio).

### PacBio amplicon analysis

The high-fidelity (HiFi) reads were assembled by circular consensus sequencing (CCS) within SMRT Link (PacBio), demultiplexed with Lima, and aligned to the reference genome GRCh38 with minimap2. The VNTR6-1 amplicons had an average read coverage of ∼10,000 reads per sample. The resulting BAM files were scored for rs56345976 and rs33961405 using SAMtools with mpileup flag (v1.17) and for VNTR6-1 using Straglr (v1.4). The analysis was restricted by reads fully covering the amplicon (GRCh38, chr5:1275500-1277500), excluding outputs from partial reads using SAMtools with the ampliconclip flag (v1.17). Phased haplotypes of rs56345976 and rs33961405 were constructed based on PacBio reads.

### DNA genotyping

TaqMan genotyping assays for *TERT* SNPs rs56345976 (C 88595060_10), rs33961405 (C 34209972_10), rs10069690 (C 30322061_10), rs2242652 (C 16174622_20), rs7705526 (C 189441058_10), rs2736100 (C 1844009_10) and rs2853677 (C 1844008_10) were purchased from Thermo Fisher. The samples were genotyped in 384-well plates on a QuantStudio 7 Flex Real-Time PCR System (Applied Biosystems) using 2x TaqMan Genotyping Master Mix (Thermo Fisher) in 5-µL reactions with 4 ng of genomic DNA per reaction.

### Analyses in the 1000 Genomes Project

High-coverage (30x) short-read whole-genome sequencing (WGS) data in CRAM format and phased genetic variants for 3,201 individuals from the 1000G populations^6^ were downloaded from https://www.internationalgenome.org/data-portal/data-collection/30x-grch38 for the 400 kb genomic region (GRCh38 chr5:1,100,000-1,500,000). The depth of coverage of aligned short-sequencing reads within the 2,290 bp genomic region corresponding to VNTR6-1 (GRCh38 chr5:1,275,210-1,277,500) was analyzed by calculating median coverage within consecutive 50-base windows using Mosdepth (v0.2.5). All the samples were classified into VNTR6-1-Short/Short genotypes (24-27 copies) and Long/any genotypes (with one or two Long alleles of 40.5 or 66.5 copies) by applying a machine learning strategy with the tidymodels framework and based on the R package ‘glmnet’ (v4.1-7)^7^. First, a total of 605 samples (18.89%) were randomly selected from the set, representing all 1000G super-populations, and visually examined and assigned to the Short or Long groups based on the coverage profiles in IGV. Then, the dataset was split into training (60%) and testing (40%) sets. Fivefold cross-validation was used during the training process to develop and evaluate the prediction model. The model demonstrated stable performance in accurately classifying VNTR6-1 into the Short and Long categories, with 96.8% specificity, 92.8% sensitivity, an F score of 0.95, and an area under the ROC curve (AUC) of 0.98 (**Figure S4**).

To identify variants predictive of VNTR6-1-Short/Long status, all 12,338 biallelic SNPs from the 1000G phased genetic variant data across the 400 kb genomic region (GRCh38 chr5:1,100,000-1,500,000) were extracted and filtered for MAF > 5%, resulting in 1,473 SNPs for analysis. Based on Chi-square tests, 594 of these SNPs were significantly associated with the VNTR6-1 Short and Long categories (p < 0.05). A random forest model was then applied using the R package ‘randomForest’ (v4.7-1.1) to identify the predictive value of the significant SNPs for VNTR6-1 categories, selecting the top 10% based on mean decrease in Gini scores. A total of 60 SNPs were identified as highly informative, with rs56345976 and rs33961405 showing the highest combined predictive probabilities for VNTR6-1 classification.

To map the haplotypes of rs56345976 and rs33961405 with the profile of coverage distribution across the genomic region GRCh38 chr5:1,275,210-1,277,500, we applied unsupervised hierarchical clustering using the core ‘hclust’ function in R (v4.3.0) with the Euclidean distance metric and complete linkage method. The rs56345976-A/rs33961405-G haplotypes captured the VNTR6-1 Long allele (Cohen’s Kappa coefficient of 0.78 and agreement of 0.90), while all the remaining haplotypes captured the VNTR6-1 Short allele (**Figure S4**). Phased data from our long-read sequencing, including assemblies and targeted PacBio sequencing, was used to confirm the co-segregation of rs56345976 and rs33961405 with VNTR6-1 (**Table S2**).

We created a custom 1000G reference panel that included all markers within the 400 kb genomic region (GRCh38 chr5:1,100,000-1,500,000). In this region, VNTR6-1 was used as a biallelic marker, with Short and Long alleles determined by the rs56345976/rs33961405 haplotypes at position chr5:1,275,400 (**Table S3**). To evaluate the scoring performance, the 1000G dataset (n=3,201) was randomly partitioned into two groups, which served as a reference panel (n=1,601) and a test panel (n=1,600) to perform phasing with SHAPEIT4 (v4.2.0) and imputation with IMPUTE2 (v2.3.2) with default settings. VNTR6-1 was confidently scored in all test panel samples (imputation quality score^8^, IQS = 0.98), with an overall concordance of 99.3% compared to the predetermined genotypes across the entire dataset. Population-specific concordance rates for VNTR6-1 imputation were as follows: EUR 99.7% (n=321), AMR 99.6% (n=243), AFR 99.1% (n=456), SAS 98.99% (n=299), and EAS 98.32% (n=281).

### Analyses in the Prostate, Lung, Colorectal and Ovarian (PLCO) Cancer Screening Trial

PLCO^9^ is a large population-based cohort that includes 155,000 participants enrolled between November 1993 and July 2001. The individual-level data, including genotyped and imputed variants and phenotype data, were provided by PLCO upon approved application. The dataset of individuals of European ancestry comprised 102,708 individuals, including 73,085 common cancer-free controls and 29,623 patients with 16 cancer types. All the variants within the 400 kb region (GRCh38 chr5:1,100,000-1,500,000) were phased using SHAPEIT4 (v4.2.0) and then VNTR6-1 genotypes (Short or Long) were assigned based on phased rs56345976/rs33961405 haplotypes. Logistic regression analyses were conducted with the logit link function for binary outcomes using the ‘glm’ function in R (v4.3.0), adjusting for sex and age.

### Analyses in the UK BioBank

Associations between genetic markers and relative telomere length (rTL) in peripheral blood lymphocytes were assessed in the UK Biobank (UKB) (https://www.ukbiobank.ac.uk/), a population-based prospective study in the United Kingdom^10^. The analysis included 351,634 cancer-free participants of European ancestry with both SNP data and rTL measurements. VNTR6-1 was scored as described above for PLCO. We used linear regression models to assess the association between the technically adjusted rLTLs (log_e_ and Z-transformed)^11^ and the genetic markers. This analysis was performed using the ‘lm’ function in R (v4.3.0) and adjusting for sex, age, and smoking status. A conditional linear model was tested by independently adding SNPs (rs2736100, rs2853677, and rs7705526) that are strongly associated with telomere length in multiple populations. To account for trend differences in rLTLs across all ages, the conditional linear model included an interaction term between the genetic markers and 5-year age groups, that were used to avoid age-heaping bias while maintaining a sufficient sample size for each age class.

### Analyses in The Cancer Genome Atlas (TCGA)

Blood-derived germline data for 9,610 TCGA participants across 33 cancer types were accessed through the National Cancer Institute Genomic Data Commons (GDC, https://gdc.cancer.gov/). Controlled access genotype calls generated from Affymetrix SNP6.0 array intensities using BIRDSUITE^12^ were retrieved from the genomic region GRCh37, chr5:335,889-2,321,650. In this region, in addition to the 5,453 initially genotyped variants, we imputed approximately 57,000 additional variants with imputation quality scores exceeding 0.8 using the TOPMed Imputation Server, which includes data from more than 97,000 participants^13^. The imputation quality scores across cancer types were as follows: mean (min–max) r^2^=0.83 (0.78-0.89) for rs56345976, r^2^=0.85 (0.75-0.92) for rs33961405, r^2^=0.85 (0.76-0.94) for rs10069690, and r^2^=0.84 (0.74-0.92) for rs2242652. Direct genotyping from germline WGS files for 387 BLCA downloaded from GDC showed high concordance rates between imputed and WGS-genotyped markers: 89.90% for rs56345976, 86.79% for rs33961405, 91.17% for rs10069690 and 92.75% for rs2242652.

Transcripts per million (TPM) for bulk *TERT* RNA-seq data were downloaded from the GDC within the Pan-Cancer Atlas publications^14^. The TPMs for the *TERT*-β and *TERT*-FL transcripts were downloaded from the UCSC Xena platform (https://xenabrowser.net/datapages/) within the UCSC toil RNA-seq Recompute Compendium, cohort TCGA Pan-Cancer (PANCAN). We used pre-computed telomerase-related metrics, including the expression-based telomerase enzymatic activity detection (EXTEND) scores based on a 13-gene signature^15^, stemness indices calculated via a predictive model using one-class logistic regression on mRNA expression^16^, a telomerase signature score estimated from a 43-gene panel, and the telomere length scores calculated using TelSeq based on WGS^17^.

eQTL analysis was conducted using TPMs for bulk RNA-seq *TERT* expression data and genetic markers (additive genetic model) using the ‘lm’ function in R (v4.3.0), with adjustments for sex and age. Spearman rank correlations between *TERT* expression (*TERT*-β and *TERT*-FL) and telomerase-associated metrics for each cancer type were determined using the ‘rcorr’ function of the Hmisc package in R (v4.3.0).

### Analyses in the Genotype-Tissue Expression (GTEx) project

TPMs for the *TERT*-β and *TERT*-FL transcripts were downloaded from the GTEx Portal (https://gtexportal.org/home/downloads/) within the bulk tissue expression database, GTEx Analysis V8 RNA-seq. Pre-computed EXTEND scores based on a 13-gene signature were obtained from the Supplementary Information of the corresponding publication^29^. Spearman rank correlations between *TERT* expression (*TERT*-β and *TERT*-FL) and EXTEND scores for each tissue type were determined using the ‘rcorr’ function of the Hmisc package in R (v4.3.0). The eQTLs for rs10069690, rs2242652 and *TERT* expression were assessed through the GTEx portal.

### CFSE proliferation assay

For each condition, cells (9.6×10^5^) were stained with a 5 µM solution of carboxyfluorescein succinimidyl ester (CFSE) dye (CellTrace CFSE Cell Proliferation Kit, Thermo Fisher) for 15 min at 37°C. Culture media containing 10% CS FBS was added to an equal volume of staining solution to quench excess dye. CFSE-stained cells (1.2×10^5^) were seeded into each well of a 6-well plate in CS serum medium and incubated at 37°C and 5% CO_2_. The remaining CFSE-stained cells were analyzed on an AttuneNxT (Thermo Fisher) flow cytometer to determine the day 0 (maximal) CFSE intensity. Seeded cells were grown for 48 h in CS serum medium to allow all cell lines to reach a sufficient level of attachment for a medium change and then switched to either full serum or CS serum medium. Forty-eight hours after the media were changed, the cells were harvested with 0.05% trypsin-EDTA and analyzed by flow cytometry to determine the final CFSE intensities. The data were analyzed using FlowJo v10. The CFSE mean fluorescence intensity (MFI) was determined by taking the geometric mean of fluorescence (collected on the BL1 channel, 530/30 nm) after gating live single cells.

### CRISPR/Cas9 genome editing

CRISPR/Cas9 guide RNAs flanking the VNTR6-1 region (1,267 bp in the reference genome) were designed using sgRNA Scorer 2.0^18^. Annealed oligonucleotides corresponding to two guide RNAs (**Table S20**) were cloned using Golden Gate Assembly cloning into PDG458 (ref ^19^, Addgene plasmid #100900; http://n2t.net/addgene:100900; RRID:Addgene 100900, a gift from Paul Thomas). The cells (1×10^6^) were transiently transfected with CRISPR/Cas9-expressing plasmids using the Amaxa 4D nucleofection system (Lonza), a 100 µl SF cell line kit, and the CM-130 program (A549 profile settings were used for all cell lines). GFP-positive cells were enriched by FACS 48 hours post-transfection using an SH800 sorter (Sony). The enriched population was further single-cell sorted in 96-well plates to isolate pure knockout populations. Genomic DNA from the expanded clones was screened by PCR with primers VNTR6-1F and VNTR6-1R (**Table S20**). These primers generate a 2,241-bp PCR product (based on the reference genome sequence) and a 974-bp PCR product after knockout. Three independent knockout clones (V6.1-KOs) were selected for functional analyses. Clones that were exposed to CRISPR reagents but did not result in knockout were compared with parental controls (WT, no CRISPR treatment) by RNA-seq analysis. CRISPR treatment had negligible effects on gene expression, and statistical analysis of RNA-seq data was performed comparing V6.1-KOs to WT.

### Cloning

The pCMV-TERT-FL-HA expression construct was generated with high-fidelity Q5 polymerase (NEB), starting from the plasmid for *TERT*-FL (GenScript OHu25394), using a forward primer with an <u>AgeI</u> recognition site and a reverse primer with HA-tag and <u>BsrGI</u> recognition sites (**Table S20**). PCR fragments were isolated by electrophoresis and a gel extraction kit (Qiagen) and cloned into an mEGFP-N1 expression vector (Addgene #54767) using AgeI and BsrGI restriction enzymes (NEB), replacing mEGFP. The pCMV-TERT-β−3xFLAG expression construct was generated using two separate Q5 PCRs from the same *TERT*-FL plasmid. The first PCR utilized the same AgeI forward primer and a reverse primer with a native <u>BamHI</u> recognition site within *hTERT* exon 9. The second PCR utilized the <u>BamHI</u> site in its forward primer and a reverse primer with 3xFLAG-tag and a <u>BsrGI</u> recognition site (**Table S20**). These two PCR fragments were isolated by electrophoresis and a gel extraction kit (Qiagen), cloned into pCR4 Blunt-TOPO (Invitrogen), and subcloned into the mEGFP-N1 expression vector using AgeI+BamHI and BamHI+BsrGI, replacing mEGFP.

### RNA extraction

Cell lysates were harvested from culture plates using 350 µl of RLT lysis buffer/well and stored at −80°C before extraction. RNA was extracted with the Qiagen RNeasy Mini RNA kit using QIAcube with standard on-column DNAse treatment (Qiagen). RNA concentrations were quantified with a Qubit RNA High Sensitivity Kit (Invitrogen).

### cDNA synthesis

7.5 µg of RNA from each sample was used in 20 µl reactions with the iScript Advanced cDNA Synthesis Kit (Bio-Rad). The cDNA was concentrated overnight by ethanol precipitation and resuspended in 37.5 µl of water, resulting in an RNA input concentration of 200 ng/µl.

### Expression assays

Expression of the *TERT*-β and *TERT*-FL transcripts was quantified with two custom TaqMan gene expression assays (Thermo Fisher, **Table S20**) designed to target specific exons and splice junctions. Reactions were multiplexed to include both targets and a custom human *HPRT1* endogenous control (NED/MGB probe, primer limited, Assay ID: Hs99999909_m1, Thermo Fisher). TaqMan reactions were run in technical quadruplicate in 384-well plates on a QuantStudio 7 Flex Real-Time PCR System (Applied Biosystems). Each 6 µl reaction included 2 µl of cDNA diluted to 100 ng/µl from a 200 ng/µl RNA input. All assays (individually and in multiplexed reactions) were validated using the *TERT*-FL-HA and *TERT*-β−3xFLAG plasmids in a 5x 10-fold dilution series (from 100 pM to 10 fM). All assays had experimentally determined PCR efficiencies of 72-100%. The identities of the PCR products were confirmed by cloning into TOPO-pCR4 vector (Invitrogen) and Sanger sequenced using M13_TOPO primers (**Table S20).**

SYBR Green RT‒qPCR assays were performed with iTaq Universal SYBR Green Supermix (Bio-Rad). The samples were run in 5 µl reactions with 2 µl of cDNA diluted to 50 ng/µl from the RNA input in 12 technical replicates on a QuantStudio 7 Flex Real-Time PCR System. The primers (10 mM, Thermo Fisher) used were identical to those used in the TaqMan assays. *HPRT1* controls (**Table S20**) were run in parallel reactions. For visualization, technical replicates of selected RT‒qPCR products were pooled and resolved on 2% agarose gels, along with a low-molecular-weight DNA ladder (NEB). Gel images were captured on a Bio-Rad ChemiDoc Imaging System and analyzed using Image Lab Software v6.1.0 (Bio-Rad). The ratios of *TERT* isoforms were calculated based on gel densitometry of the PCR products (120 bp and 302 bp).

Total *TERT* expression was measured in 5 µL reactions using TaqMan assays (FAM, exons 3-4) with *TERT*-Hs00972650_m1 multiplexed with the endogenous control *HPRT1* (VIC, primer-limited, Assay ID: Hs99999909_m1) and TaqMan Gene Expression Buffer (all from Thermo Fisher).

### RNA-seq

RNA quality (all RINs>9.0) was verified using the Bioanalyzer (Agilent) and an RNA 6000 Nano Kit (Agilent). For each sample, 200 ng of total RNA was used to prepare an adapter-ligated library with the KAPA RNA HyperPrep kit with RiboErase (HMR) (KAPA Biosystems) using xGen Dual Index UMI Adapters (IDT). The multiplexed libraries with 250-350 bp inserts were sequenced on a NovaSeq 6000 (Illumina) to generate 279 to 418 million paired-end 150 bp reads per sample. Quality assessment of RNA-seq data was conducted using MultiQC (v1.16)^20^. Quantification of transcript abundance was performed using Salmon (v0.14.1) in count mode with —validateMappings flag and expressed as transcripts per million (TPM). The raw RNA-sequencing reads were aligned with STAR^21^ based on the reference genome GRCh38 and GENCODE annotation (v36). Differential expression analysis was conducted with DESeq2 (v1.40.2) based on the estimated counts obtained from Salmon quantification, controlling for the false discovery rate (FDR). Gene-level transcript abundances were estimated with ‘lengthScaledTPM’ in the R package ‘tximport’ (v1.28.0). Gene Ontology (GO) analysis and gene set enrichment analysis (GSEA) on differentially expressed genes was conducted with clusterProfiler (v4.8.3).

### G4 Hunter prediction analysis

Analysis was performed with G4Hunter (https://bioinformatics.ibp.cz)^22^. PacBio-generated DNA sequences for UMUC3 (24 repeat copies per each allele) and HG03516 (27 and 66.5 repeat copies per allele) were used as inputs flanked by 120 bp on each side of the V6.1 region.

### G4-seq analysis

For the lymphoblastoid cell line NA18057 (VNTR6-1-Short/Short genotype, 24 and 27 repeat copies), ChIP-seq data for G quadruplexes (G4) detected in forward and reverse orientations were downloaded from BED files from the GEO dataset GSE63874 (ref ^23^, files GSE63874_Na_K_minus_hits_intersect.bed.gz and GSE63874_Na_K_plus_hits_intersect.bed.gz). These files were merged into a single BED file and converted to the UCSC BED format. The G4 mismatch quantification bedGraph files GSE63874_Na_K_12_minus.bedGraph.gz and GSE63874_Na_K_12_plus.bedGraph.gz were downloaded and converted into bigwig format using the bedGraphToBigWig tool.

Similarly, for the 293T normal embryonic kidney cell line (V6.1-Long/Long genotype), the G4-seq data were downloaded from GSE110582 (ref ^24^, files GSM3003539_Homo_all_w15_th-1_minus.hits.max.K.w50.25.bed.gz and GSM3003539_Homo_all_w15_th-1_plus.hits.max.K.w50.25.bed.gz) and processed as above. The G4 mismatch quantification values were downloaded from GSM3003539_Homo_all_w15_th-1_minus.K.bedGraph.gz and GSM3003539_Homo_all_w15_th-1_plus.K.bedGraph.gz. The G4-seq tracks for NA18057 and 293T cells were visualized through the UCSC Genome Browser (GRCh37).

### Evaluation of G4 ligands

Five G4 stabilizing ligands were tested for their ability to stabilize TERT G4. Ligands: PhenDC3, TMPyP4, BRACO-19 and Pyridostatin were provided by Dr. John Schneekloth. Pidnarulex (CX5461) was selected from the literature^25^ and obtained from Selleck Chem. For optimization, UMUC3 cells (4×10^5^) were seeded into each well of a 6-well plate. After adhering for 24 hours, the cells were treated for 24, 48, or 72 hours with ligands at 0.1 µM, 0.3 µM, 1 µM, 3 µM, 10 µM, or 30 µM dissolved in DMSO, with DMSO vehicle alone and untreated control samples included on each plate. In the 72-hour group, the media was replaced at 48 hours, and the cells were harvested at 72 hours. The viability of the treated cells was evaluated by cell counting with a Lionheart FX automated microscope (Agilent) every 24 hours. Pidnarulex (CX5461) and PhenDC3 at 3 µM for 72 hours were identified as the most effective treatments for modulating *TERT* exon 7-8 skipping and were used in subsequent experiments. UMUC3 WT and V6.1 KO cells were treated in technical replicates in three independent experiments.

### Western blot

BCA-normalized protein samples and 10 µL of SeeBlue Plus2 ladder were loaded and run on gels using 1X Bolt running buffer at 165 V for 1 hour and transferred to nitrocellulose membranes using an iBlot2 dry transfer instrument (Invitrogen). The membranes were blocked with 5% milk in 1X TBST for 1 hour at room temperature. The membranes were incubated overnight at 4°C with primary antibodies in 2.5% milk in 1X TBST (anti-GFP: Invitrogen A-11122; anti-HA: Novus NB600-362; anti-FLAG: Sigma M2; anti-GAPDH: Abcam ab9485). After three 5-min washes with 1X TBST, the membranes were incubated at room temperature for 1 hour with secondary antibodies (anti-rabbit: Cell Signaling 7074; anti-mouse: Cell Signaling 7076; anti-goat: Santa Cruz sc-2304) and imaged using Pico and Femto ECL reagents (Thermo).

### Structured illumination microscopy fluorescence imaging

A549 cells were chosen for imaging of mitochondria because this highly transfectable cell line has a larger cytoplasmic area than UMUC3, allowing better visualization. The cells were seeded in a 12-well plate at 1.25×10^5^ cells/ml and co-transfected with pCMV-TERT-FL-HA or pCMV-TERT-β−3xFLAG expression constructs at a 50:50% isoform ratios. Transfections were performed using Lipofectamine 3000 for 4 hrs. Transfected cells were washed with DPBS, dissociated using Accutase (StemPro), and counted. The cells were then diluted and seeded onto CultureWell Chambered Coverglass (Intigrogen). After 48 hours, the coverslips were fixed with 4% formaldehyde in PBS for 10 min, permeabilized with 0.03% Triton-X 100 for 10 min, and blocked with blocking buffer (5% BSA + 0.01% Triton-X 100 in PBS) for 30 min. The coverslips were incubated at 4°C overnight with the following primary antibodies: anti-FLAG (Sigma M2, mouse, 1:400 dilution), anti-HA (Novus NB600-362, goat, 1:400 dilution), and anti-TOM20 (Proteintech 11802-1-AP, rabbit, 1:1000 dilution) diluted in blocking buffer, followed by incubation at room temperature for 30 min with the following secondary antibodies: anti-mouse-AlexaFluor488 (Thermo Fisher A21202, 1:500 dilution), anti-goat-AlexaFluor647 (Thermo Fisher A32849, 1:500 dilution), and anti-rabbit-AlexaFluor555 (Thermo Fisher A31572, 1:500 dilution) diluted in blocking buffer. Three washes were performed with PBS between all staining steps; after the final wash, the cells were counterstained with 3 µg/ml DAPI. The coverslips were then mounted onto glass slides with ProLong Gold Antifade Mountant (Invitrogen) and sealed with clear nail polish. Super-resolution structured illumination microscopy fluorescence images were obtained using ZEN Black software on an ELYRA PS.1 Super Resolution (SR) microscope (Carl Zeiss, Inc.) with a Plan-Achromat 63X/1.4 NA oil objective and a Pco.edge sCMOS camera, 405 nm/488 nm/561 nm/633 nm laser illumination and standard excitation and emission filter sets. Raw data were acquired by projecting grids onto the sample generated from the interference from a phase grating with 23 µm, 28 µm, and 34 µm spacings for 405 nm, 488 nm and 561 nm excitation, respectively (3 grid rotations and 5 grid shifts for a total of 15 images per super-resolved z-plane per color). The raw images were processed with ZEN black software. For publication, images were scaled to 8-bit RGB identically with a linear LUT and exported in TIFF format using ImageJ. Figures were made from the TIFF images in Adobe Illustrator without any change in resolution, except for the inset zoomed images.

### Apoptosis assay

Cells (1.2×10^5^) were seeded in each well of 6-well plates (Corning), and the media was changed 48 h later to complete serum, CS serum medium or medium supplemented with 10 µM cisplatin. Cells were harvested with 0.05% trypsin-EDTA 48 h after media change, pelleted at 500 ×g for 5 min, and washed with 1 mL of PBS. The cells were stained with an Annexin V-FITC conjugate (Thermo Fisher) and propidium iodide (Thermo Fisher) in Annexin V staining buffer (Thermo Fisher) according to Rieger et al^26^. FITC (ex.488 nm/em.517 nm) and PI (ex.488 nm/em. 617 nm) fluorescence were analyzed by flow cytometry on an Attune NxT with a CytKick Autosampler (Thermo Fisher). Unstained cells, Annexin V-FITC-stained cells, and PI-stained UMUC3 cells were used as compensation controls. Apoptosis was determined by the percentage of FITC-positive cells.

### Cell density analysis

UMUC3 and three V6.1-KO clones were seeded in 6-well plates (Falcon) at 4×10^4^ cells/well in EMEM. After adherence for 24 hours, the cells were grown for 72, 96, 120, or 144 hours, with one time point per plate. The cells were washed with 1 mL of PBS and harvested on plates with RLT lysis buffer (Qiagen). RNA was extracted using the QiaCube RNeasy Mini protocol (Qiagen), followed by cDNA preparation, *TERT* qPCR, and gel densitometry as described above.

### xCELLigence Real-Time Cell Analysis (RTCA)

For RTCA, 5637 cells were seeded in a 12-well plate at 1.25×10^5^ cells/ml and transfected with either GFP, pCMV-TERT-FL-HA, or pCMV-TERT-β−3xFLAG expression constructs either as single transfection or co-transfection at different ratios of isoforms (80:20%, 50:50% and 20:80%). Transfections were performed using Lipofectamine 3000 for 4 hrs. Transfected cells were washed with DPBS, dissociated using Accutase (StemPro), and counted. The cells were then diluted and seeded into an xCELLigence E-Plate 16 microplate (Agilent) at 1.0×10^3^ cells/well and placed on an xCELLigence RTCA DP system (Agilent). The data were collected every 15 minutes in RTCA software for 288 hours and then exported for analysis.

In a separate experiment, 1.0×10^3^ UMUC3-WT or UMUC3-V6.1-KO cells grown in CS serum medium were seeded into each well of an E-Plate 16 (Agilent). Cell label-free impedance in the E-Plate (correlated with cell proliferation) was measured every 15 minutes for 283 hours using an xCELLigence RTCA DP system. Two days after seeding, the medium was changed to either full serum medium or CS serum medium (control).

Linear mixed models were applied to the impedance data obtained from the xCELLigence system, where the treatment type was considered a fixed effect term and the technical replicate was considered a random effect term. Maximum likelihood estimation procedures were employed to conduct joint effects likelihood-ratio tests, while restricted maximum likelihood estimation was utilized for more precise estimation of effect sizes as beta coefficients using the linear mixed-effects function in the R package ‘nlme’ (v3.1–162).

### HiChIP analysis

The H3K27Ac HiChIP libraries for the bladder cancer cell lines T24 and RT4 were generated using the Arima-HiChIP protocol (Arima Genomics, A101020). Briefly, 1×10^6^ cells/replicate were collected for chromatin cross-linking followed by digestion with a restriction enzyme cocktail, biotin labeling, and ligation. The samples were then purified, fragmented, and enriched. Pulldown was performed using an antibody against H3K27ac (Cell Signaling, #8173). The Arima-HiChIP libraries that passed QC were sequenced using an Illumina NovaSeq 6000 to generate raw FASTQ files for each sample. The paired-end reads were aligned to the GRCh37 genome using the HiC-Pro pipeline (v3.1.0, https://github.com/nservant/HiC-Pro). The confirmed interaction reads were used as input for significant loop calling via the FitHiChIP tool (v.11.0, https://github.com/ay-lab/FitHiChIP) with default settings. The HiChIP loop and ATAC peak calling files for the GM12878 and normal bladder samples were downloaded from the Gene Expression Omnibus (GSE188401). The interactions were visualized through the UCSC genome browser.

### PacBio DNA methylation analysis

Freshly collected genomic DNA (5 µg) from the HT1376, RT4, T24, SCaBER, UMUC3, and Raji cell lines was sheared using Covaris g-tubes at 4800 rpm, followed by size selection using PippinHT. Three SMRT flow cells were run for each sample library on the PacBio Sequel II platform. The sequence reads were transformed into FASTQ and aligned to the GRCh38 reference genome using the default settings of the SMRT-Link workflow. 5mC DNA methylation analysis was a part of the SMRT-Link pipeline, and the corresponding information specifying the positions and probabilities of 5mC methylation at CpG sites was integrated into the output file.

### Oxford Nanopore cDNA-seq

cDNA libraries were generated using the PCR cDNA Sequencing Kit SQK-DCS109 (Oxford Nanopore Technologies), starting with 100 ng of poly-A RNA. Libraries were loaded onto R9.4.1 PromethION flow cells mounted on a P2 Solo and run for 96 hours. Basecalling was performed using MinKNOW software with the high-accuracy model on a GridION sequencer (Oxford Nanopore Technologies). Reads were aligned to GRCh38 via Minimap2 (v2.26) and SAMtools (v1.5). UMUC3 yielded 25,827,200 reads, with 46 reads aligning to *TERT*, whereas UMUC3 V6.1 KO yielded 18,709,848 reads, with 62 reads aligning to *TERT*.

### Analysis of sequence conservation in non-human species

Haplotype-resolved Telomere-to-Telomere (T2T) assemblies of primates were downloaded from GenomeArk (https://www.genomeark.org/). The FASTA sequences were aligned to the human GRCh38 reference genome using Minimap2 (v2.26) with the ‘-ax asm10’ flag and converted to a BAM file using SAMtools (v1.5). The *TERT* V6.1 repeat units were analyzed with Tandem Repeat Finder (https://tandem.bu.edu/trf/trf.html). The BAM files of Neandertal (n=3) and Denisova (n=1) individuals were downloaded from the Max Planck Institute for Evolutionary Anthropology resource (http://cdna.eva.mpg.de/neandertal/Vindija/bam/Pruefer_etal_2017/ and http://cdna.eva.mpg.de/neandertal/Chagyrskaya/) and visualized using IGV.

### Statistical analysis

Unless indicated, analyses were performed with R Studio (v4.3.0), GraphPad Prism (v.8) and FlowJo (v9); p values are for unpaired two-sided Student’s t tests or linear regression adjusted for the indicated covariates.

This work utilized the computational resources of the NIH HPC Biowulf cluster (http://hpc.nih.gov).

