## Supplementary Figures for "Genetic regulation of *TERT* splicing contributes to reduced or elevated cancer risk by altering cellular longevity and replicative potential"

**Florez-Vargas et al**

**Figure S1.** Analysis of the *TERT* VNTR6-1 and VNTR6-2 in long-read human genome assemblies.

**Figure S2.** VNTR6-1 in short-read sequencing alignments in the 1000G samples.

**Figure S3.** Random forest analysis of SNP-based prediction of VNTR6-1 groups in 3,201 individuals of diverse ancestries from the 1000G.

**Figure S4.** VNTR6-1 scoring in the 1000G samples.

**Figure S5.** VNTR6-1 and rs56345976/rs33961405 in targeted long-read PacBio sequencing alignments.

**Figure S6.** Linkage disequilibrium (LD) profile of the *TERT* region in the custom reference panel of 1000G-EUR populations.

**Figure S7.** Elimination of *TERT*-VNTR6-1 by CRISPR/Cas9 gene editing in UMUC3.

**Figure S8.** Elimination of *TERT*-VNTR6-1 by CRISPR/Cas9 gene editing in A549 and effects on *TERT* splicing.

**Figure S9.** DNA methylation profile of the *TERT* VNTR6-1 in PacBio whole-genome sequencing (WGS) alignments.

**Figure S10.** Lack of long-range chromatin interactions within the *TERT* region.

**Figure S11.** Analysis of G-quadruplexes (G4) formed by VNTR6-1.

**Figure S12.** Alternative splicing generates a novel *TERT*-Δ8 isoform.

**Figure S13.** Linkage disequilibrium plots ($r^2$) for individuals from 1000G-EUR and 1000G-AFR reference panels, and patients with Burkitt lymphoma (88% of African ancestry).

**Figure S14.** Analysis of *TERT* intron retention in 78 Burkitt lymphoma tumors.

**Figure S15.** Analysis of splicing events between *TERT* exons 4 and 5 in relation to GWAS leads rs10069690 and rs2242652 in 78 Burkitt lymphoma tumors.

**Figure S16.** Main *TERT* isoforms with alternative splicing within the area of GWAS leads rs10069690 and rs2242652.

**Figure S17.** VNTR6.1-associated responses to changes in environmental conditions.

**Figure S18.** Effects of overexpression of *TERT* isoforms.

**Figure S19.** Cellular localization of the TERT-FL and TERT-β protein isoforms.

**Figure S20.** Association analysis of cancer risk in PLCO and relative leukocyte telomere length (rLTL) in UKB cancer-free individuals.

**Figure S21.** Analysis of isoform-level *TERT* expression in relation to telomerase-associated metrics in GTEx and TCGA.

**Figure S22.** *TERT* VNTR6.1 region across vertebrate species.

**Figure S23.** *TERT* VNTR6-1 profiles in WGS short-read alignments in modern and archaic humans.

**Figure S24.** Age-dependent frequencies of the human-specific alleles in individuals of European ancestry.

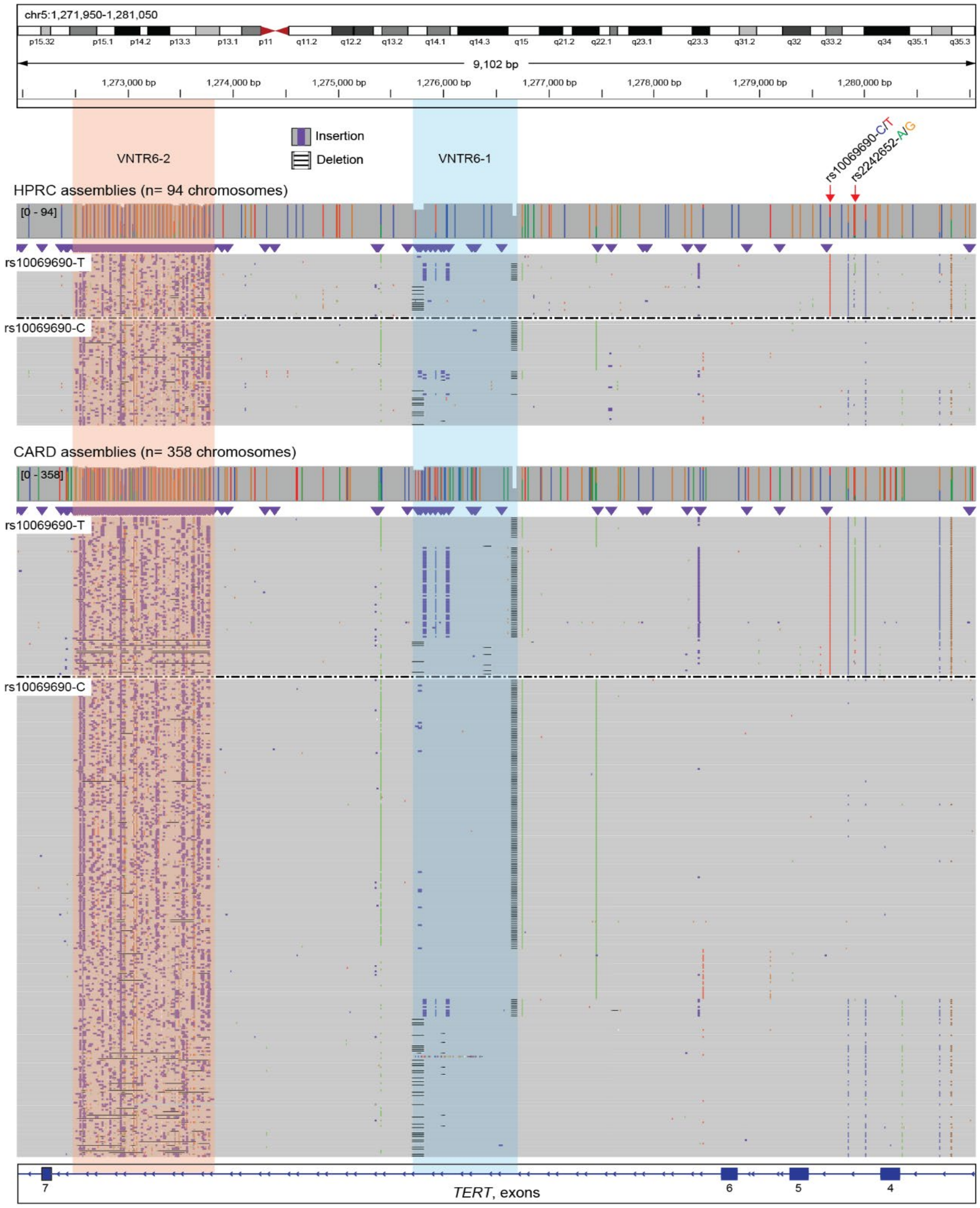


**Figure S1. Analysis of the *TERT* VNTR6-1 and VNTR6-2 in long-read human genome assemblies.**

Integrative Genomics Viewer (IGV) plots of long-read human genome assemblies (n=452) focusing on the GRCh38 chr5:1,250,000-1,450,000 region featuring VNTR6-1 (blue highlight) and VNTR6-2 (pink highlight) within intron 6 and GWAS leads rs10069690 and rs2242652 within intron 4 (gray – base matches, colored – mismatches with the reference genome). Each individual is represented by two assemblies (reads). Insertions (purple marks) in the VNTR6-1 region are enriched in the assemblies with rs10069690-T and rs2242652-A alleles. Compared with the alternative alleles, assemblies with the rs2242652-A allele have more VNTR6-1 copies (p=5.93E-19) and VNTR6-2 (p=7.66-04). Assemblies with the rs10069690-T allele have more copies of VNTR6-1 (p=5.40E-11) but not VNTR6-2 (p=0.84). Gray marks within the VNTR6-1 region correspond to the deletion of 53 bp (~1.5 repeats) segregating with 25.5, 40.5 and 66.5 VNTR6-1 alleles but independent of rs10069690 and rs2242652.

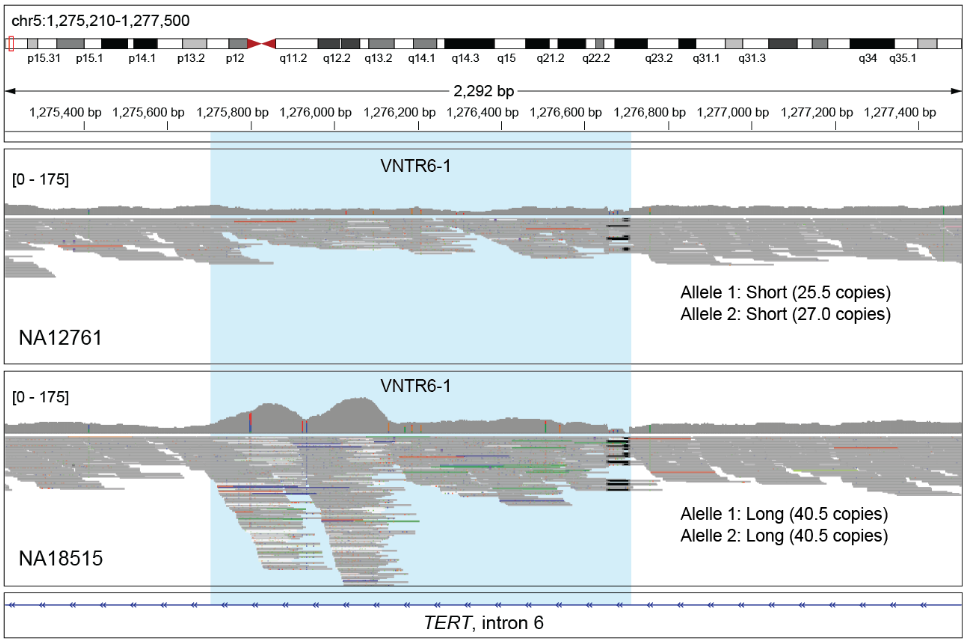


**Figure S2. VNTR6-1 in short-read sequencing alignments in the 1000G samples.**

Representative IGV plots of short-read WGS alignments (Illumina, 30x coverage) illustrating the repeat number profiles of VNTR6-1 (blue highlight) for two 1000G samples, NA12761 and NA18515. The VNTR6-1 repeat sizes were determined by targeted PacBio sequencing, identifying NA12761 as Short/Short genotype (25.5 and 27 copies) and NA18515 as Long/Long genotype (homozygous for 40.5 copies).

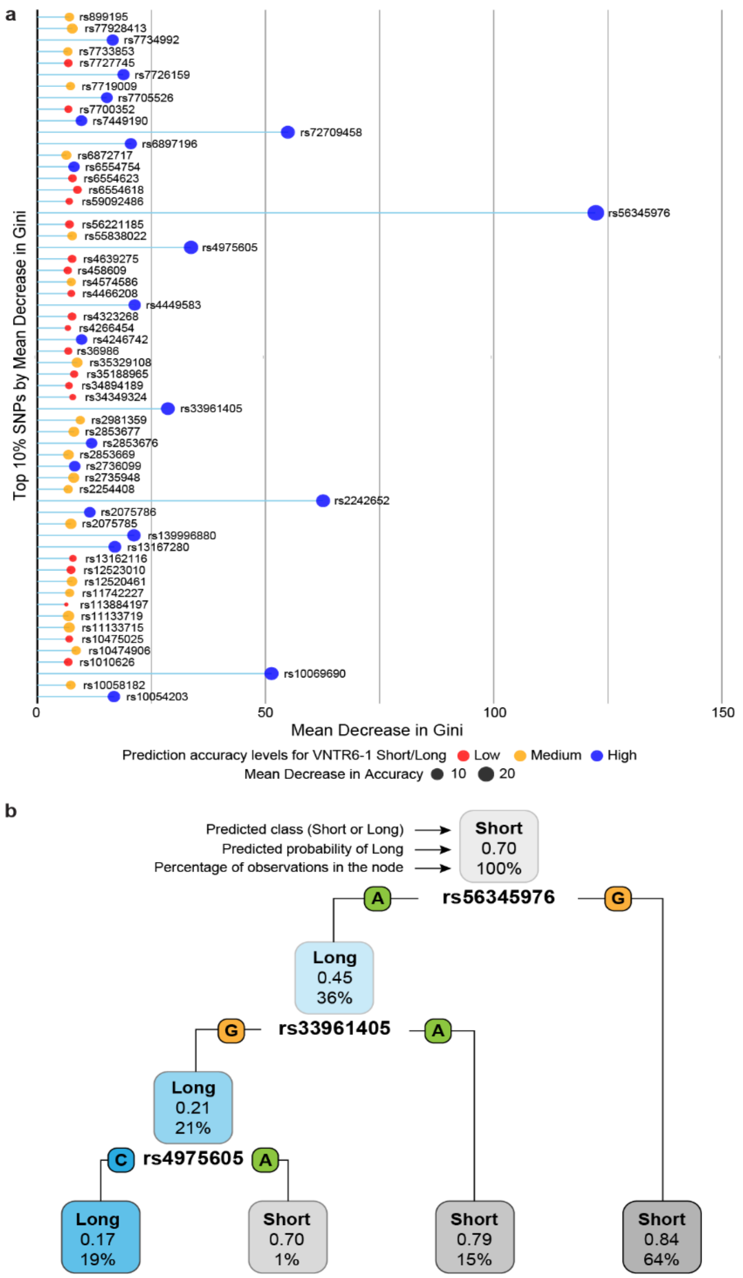


**Figure S3. Random forest machine learning analysis of SNP-based prediction of VNTR6-1 groups in 3,201 individuals of diverse ancestries from the 1000G.**

**a**, The top 10% of SNPs within the 400 kb genomic region (GRCh38 chr5:1,100,000-1,500,000) were selected based on Mean Decrease in Gini values, indicating their high discriminative power to predict Short/Short and Long/any genotype groups, which were established for each sample based on short-read sequencing depth profiles. Dot sizes represent a Mean Decrease in Accuracy, with larger circles corresponding to higher model accuracy and higher values corresponding to substantial accuracy loss if the feature is removed. SNPs with both a high Mean Decrease in Gini and a high Mean Decrease in Accuracy are the most informative for distinguishing the VNTR6-1 groups. **b**, A representative decision tree from the ensemble of 500 trees in the random forest model illustrates the classification of VNTR6-1 groups. Each node in the tree represents a decision point based on the values of the predictor variables, leading to sample assignment either to Short/Short or Long/any groups.

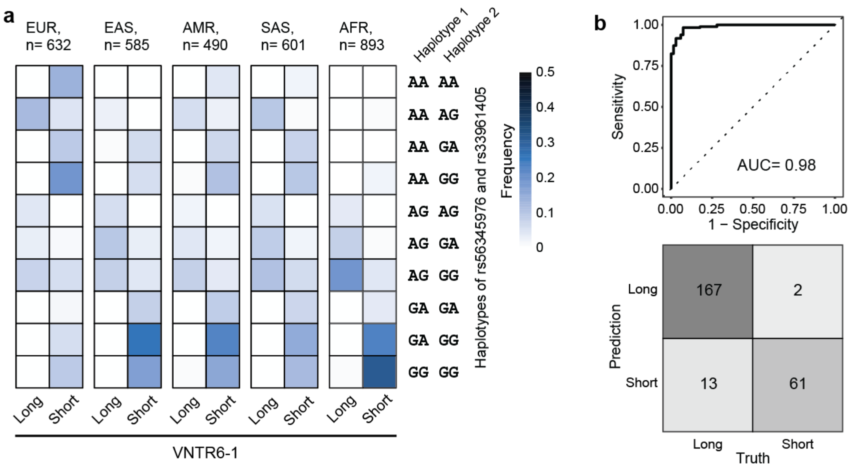


**Figure S4. Machine learning scoring of VNTR6-1 in the 1000G samples.**

**a**, Analysis in the 1000G super-populations shows that the rs56345976-A/rs33961405-G haplotype segregates with the VNTR6-1-Long group determined by short-read genomic coverage profiles, whereas all other haplotypes segregate with the VNTR6-1-Short group. **b**, The receiver operating characteristic (ROC) curve and confusion matrix results for VNTR6-1 classification into the Short and Long groups comparing assignments based on rs56345976-A/rs33961405-G haplotype to "Truth" – classification into Long/any and Short/Short genotype groups based on genomic coverage profiles.

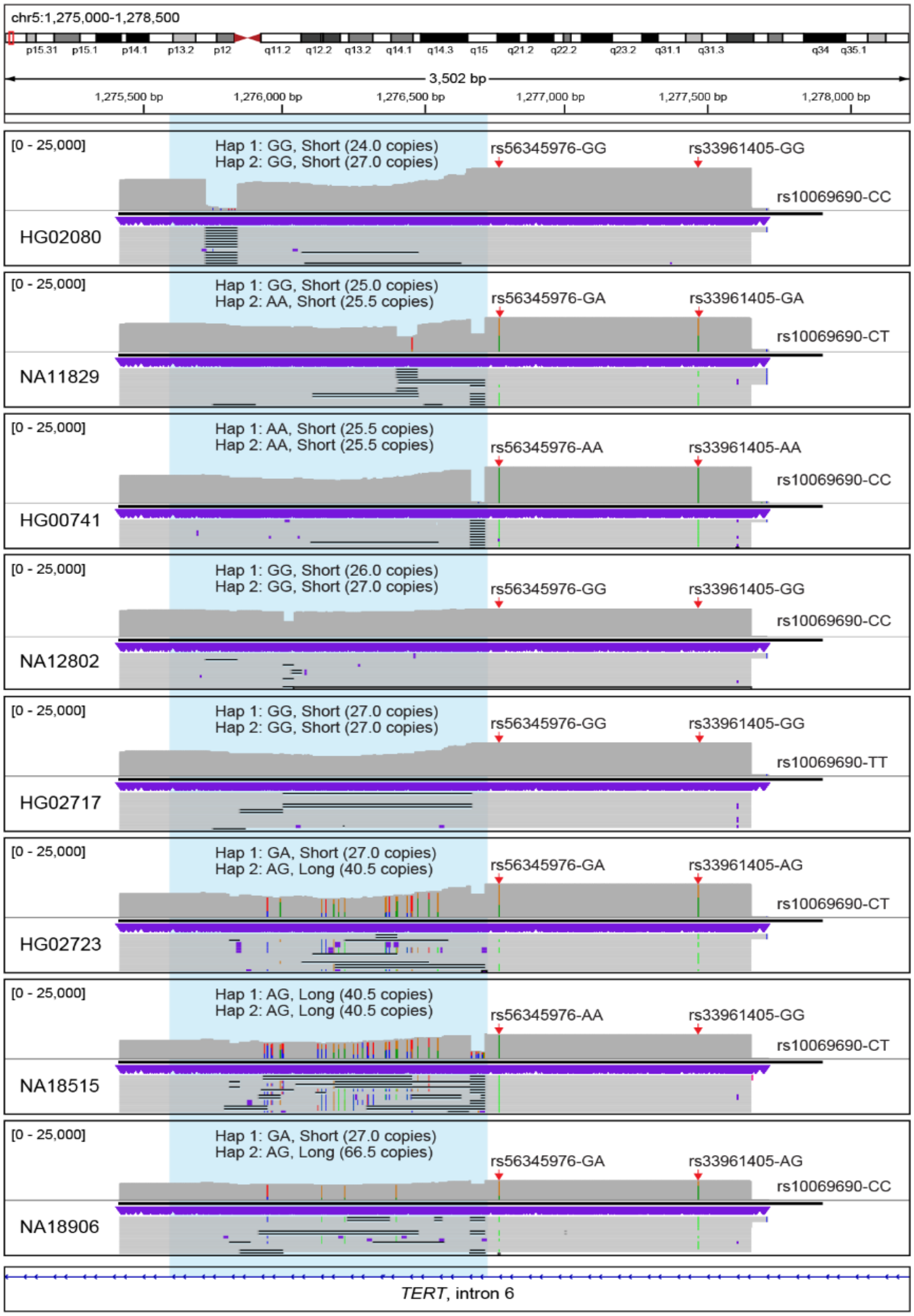


**Figure S5. VNTR6-1 and rs56345976/rs33961405 in targeted long-read PacBio sequencing alignments.**

Representative IGV plots for targeted PacBio sequencing alignments for HapMap/1000G samples illustrating VNTR6-1 (blue highlight) and SNPs rs56345976 and rs33961405 (red arrows). Hap 1 and Hap 2 present phased haplotypes of rs56345976 and rs33961405 alleles and corresponding VNTR6-1 repeat copies determined based on Tandem Repeat Finder analysis of PacBio sequencing reads and HPRC long-read genome assemblies (where available). The select samples demonstrate the range of VNTR6-1 repeat sizes observed in human populations (24-66.5 copies).

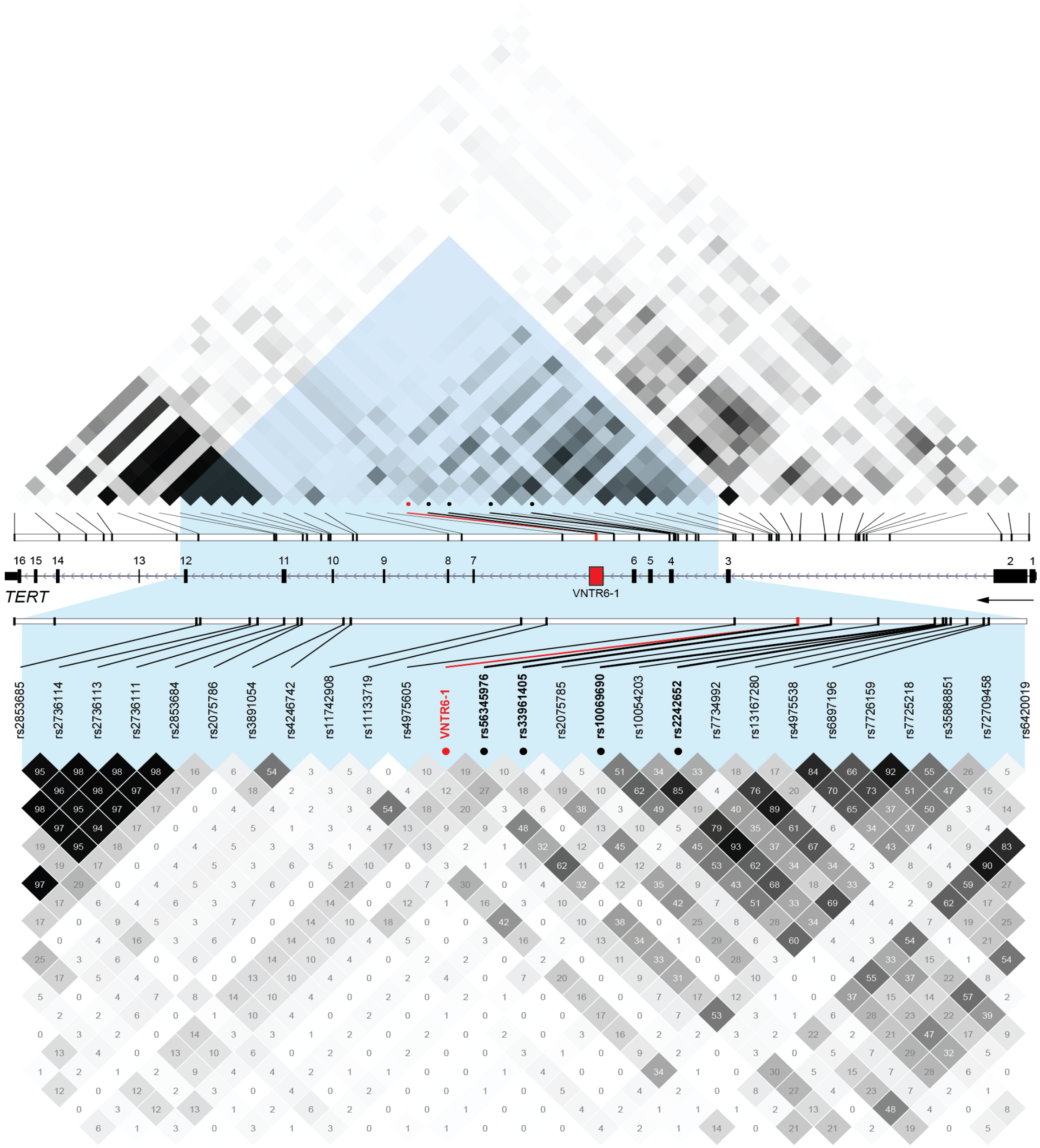


**Figure S6. Linkage disequilibrium (LD) profile of the *TERT* region in the custom reference panel of 1000G-EUR populations.**

The custom reference panel for 632 individuals of European ancestry from 1000G populations was constructed for the 400 kb genomic region GRCh38, chr5:1,100,000-1,500,000 by adding VNTR6-1 marker (Short and Long alleles) to the existing high-coverage (30x) WGS genotype data downloaded from https://www.internationalgenome.org/data-portal/data-collection/30x-grch38 (shown at MAF>0.05, HWE p>0.001). The upper panel shows the whole *TERT* region, and the lower panel details the area with the relevant markers – VNTR6-1, which is constructed based on rs56345976/rs33961405 haplotypes, and GWAS leads in intron 4 – rs10069690 and rs2242652. Darker shading on the LD plots corresponds to high $r^2$ between markers, also shown as values in boxes.

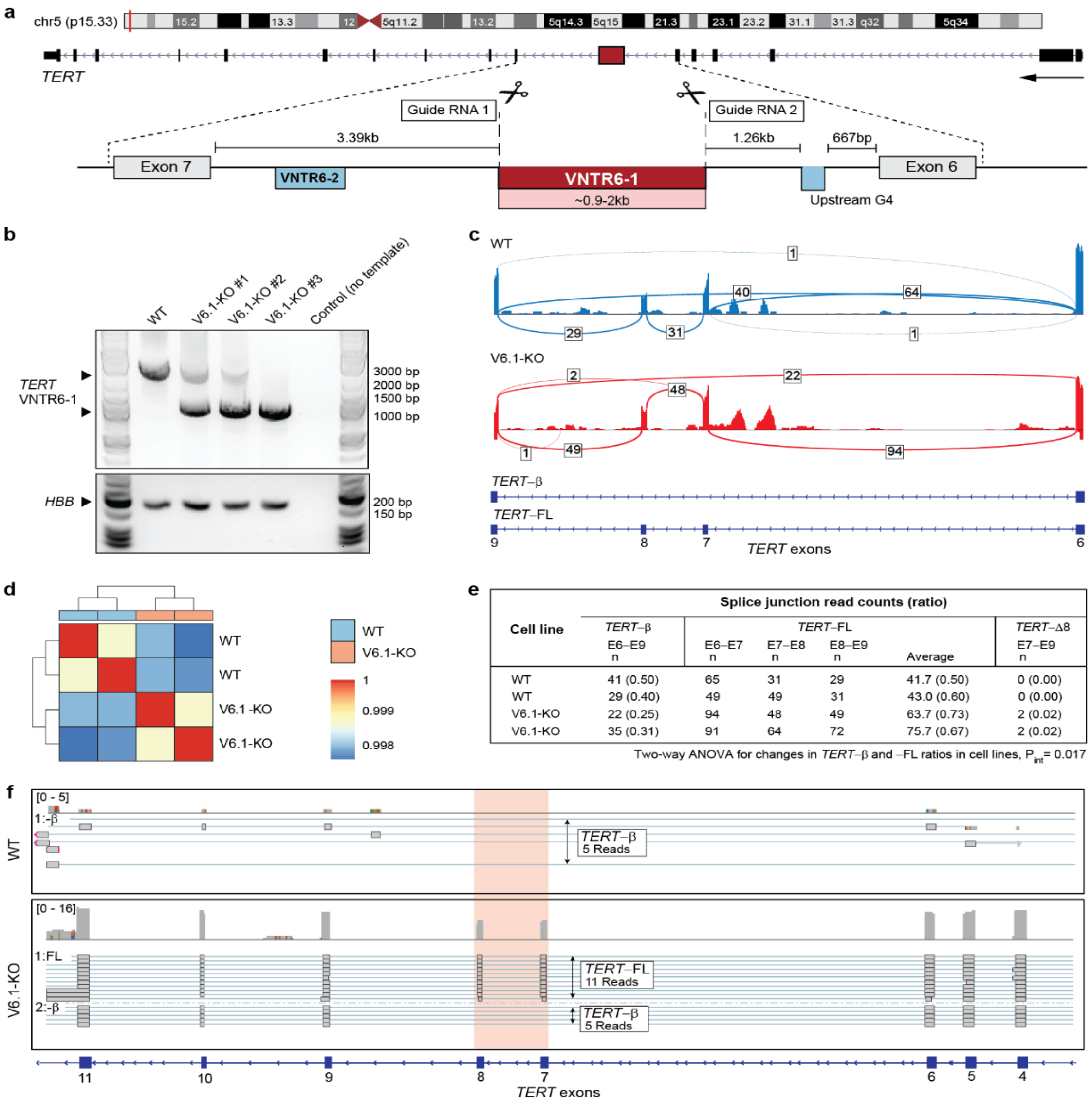


**Figure S7. Elimination of *TERT*-VNTR6-1 by CRISPR/Cas9 gene editing in UMUC3 cells.**

**a**, CRISPR/Cas9 gene editing strategy for creating VNTR6-1-KO cell lines. The 20 nt protospacers of CRISPR/Cas9 guide RNAs were placed 133 bp upstream and 82 bp downstream of VNTR6-1 within intron 6. **b**, Agarose gels of PCR products amplified from genomic DNA of UMUC3 cells; *HBB* amplicon is used as a normalization control. **c,** Representative IGV-Sashimi plots showing RNA-seq splicing profiles of WT and VNTR6-1-KO cell lines. **d,** A heatmap showing similar clustering of biological RNA-seq duplicates of WT and VNTR6-1-KO. **e,** RNA-seq splicing profiles based on biological duplicates of WT and VNTR6-1-KO cell lines (including one from plot **c**). VNTR6-1-KO causes a significant (ANOVA p=0.017) change in *TERT* splicing pattern, with an increase in *TERT*-FL and a decrease in *TERT*-β isoform expression. **f**, IGV coverage map for Oxford Nanopore cDNA sequencing of WT and VNTR6-1-KO cell lines; the highlighted region between *TERT* exons 7 and 8 shows the position of *TERT*-FL and *TERT*-β, with an increase of *TERT*-FL in VNTR6-1-KO cell line, similarly to what was observed by short-read RNA-seq (panel **c-e**).

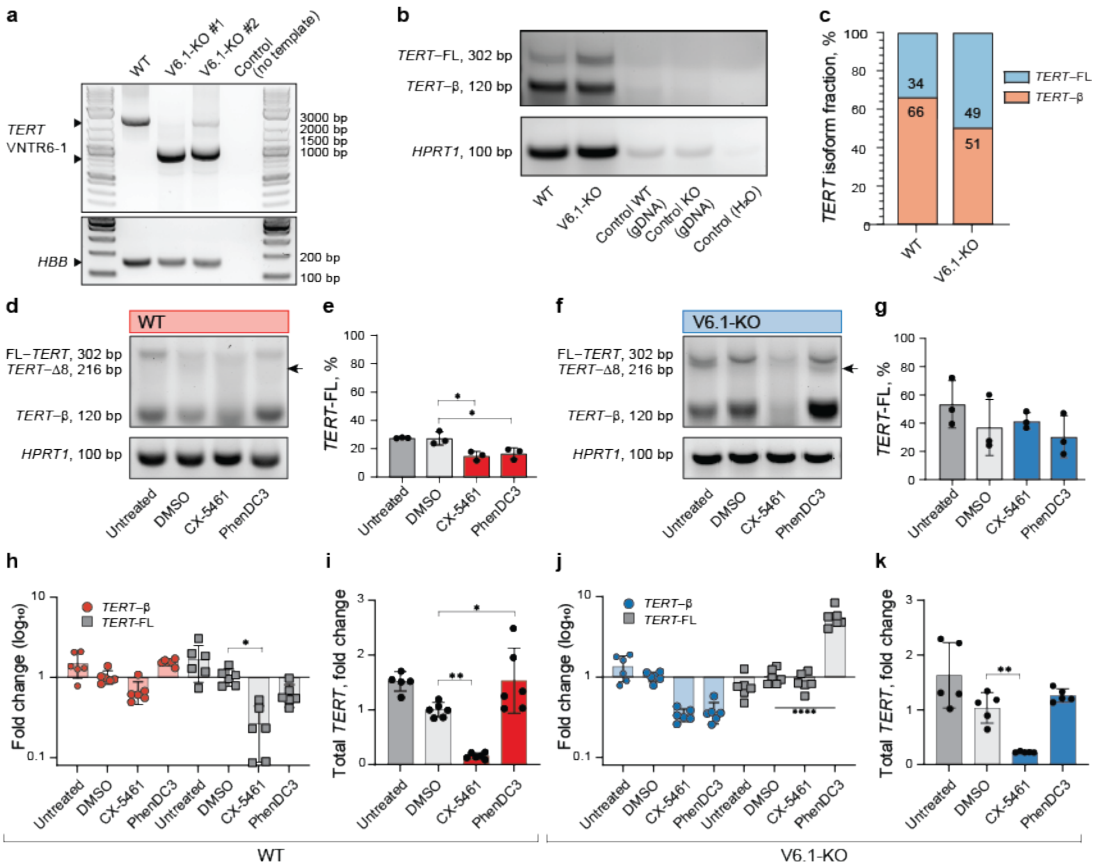


**Figure S8. Elimination of *TERT*-VNTR6-1 by CRISPR/Cas9 gene editing in A549 cells and effects on *TERT* splicing.**

**a**, Agarose gels of PCR products amplified from genomic DNA of A549; *HBB* amplicon is used as a normalization control. **b**, Agarose gels of RT–PCR products amplified from cDNA of corresponding samples; gDNA–genomic DNA was used as a negative control; *HPRT1* was used as a normalization control. **c**, Densitometry results of the PCR amplicons in plot **b**. Experiments in A549 cells comparing *TERT* splicing and isoform-specific expression after 72 hrs of treatment with G4 stabilizing ligands, normalized to *HPRT1* as an endogenous control in the WT (**d**, **e**) and V6.1-KO (**f**, **g**) cell lines. **d**, **f**, A representative agarose gel of SYBR-Green RT–qPCR products detecting several isoforms with primers located in exons 6 and 9. The extra PCR band, marked by an arrow in panels **c** and **d**, is further explored in **Figure S12**. **e**, **g**, Densitometry analysis of the corresponding agarose gels evaluating the percentage of *TERT-FL* (%) relative to the total PCR products. **h**, **j**, Isoform-specific TaqMan RT-qPCR analysis of *TERT-FL* and *TERT-β* following treatment with G4-stabilizing ligands CX-5461 and PhenDC3 for 72 hours. **i**, **k**, Total *TERT* expression measured by TaqMan RT-qPCR assay for exon 3-4. All analyses are based on one of three representative experiments. Comparisons were made against the vehicle control (DMSO). All analyses are based on three experiments, with one representative gel shown. Comparisons were made against the vehicle control (DMSO). Statistical significance is indicated as follows: **$p < 0.01$, ***$p < 0.001$, ****$p < 0.0001$, Student's t test.

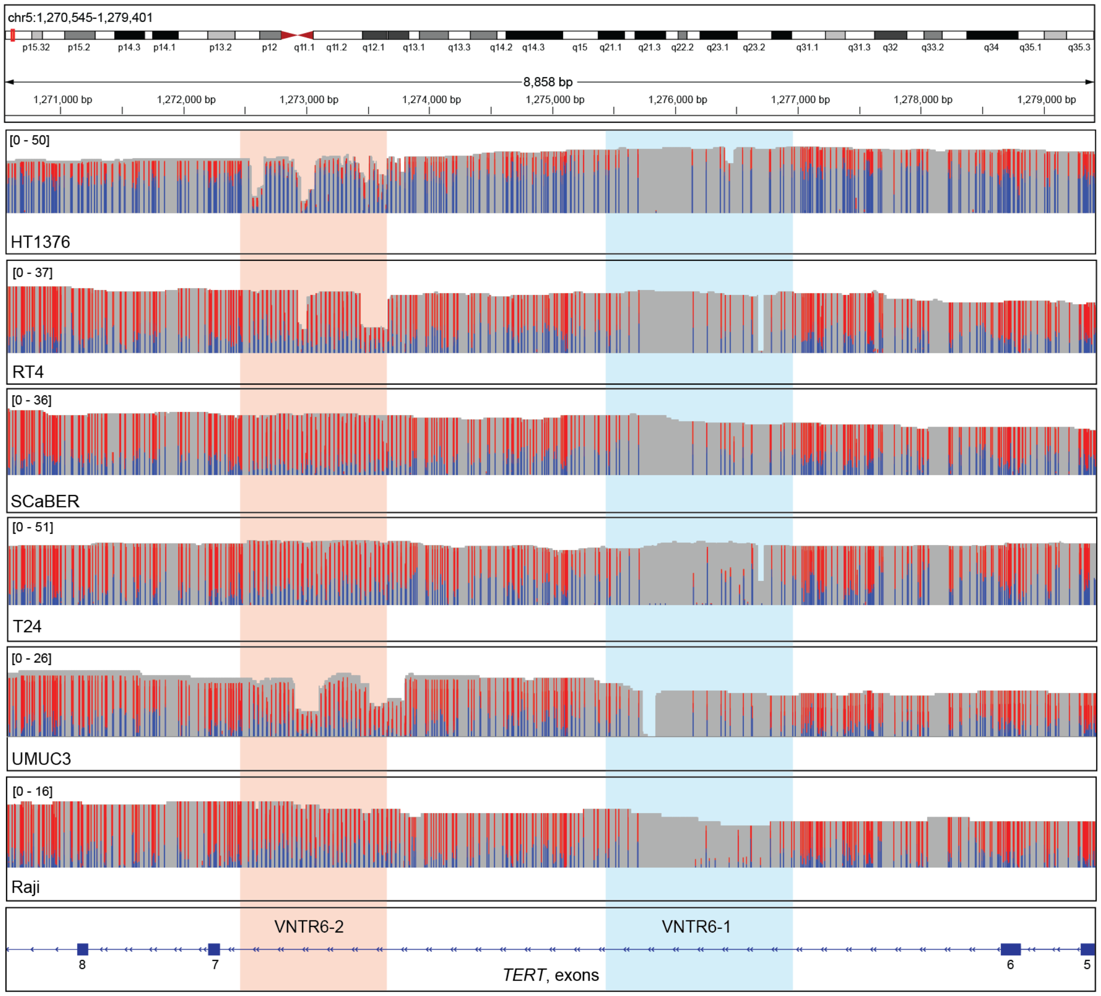


**Figure S9. DNA methylation profile of the *TERT* VNTR6-1 in PacBio whole-genome sequencing (WGS) alignments.**

IGV plots of PacBio WGS in bladder cancer cell lines HT1376, RT4, T24, SCaBER, UMUC3, and a Burkitt lymphoma cell line Raji. CpG sites are marked based on the probabilities of 5-methylcytosine modifications (5mC), with blue (< 50%) and red (≥50%); gray areas - no CpG sites. The areas of VNTR6-1 and V6.2 within *TERT* intron 6 are highlighted, with the VNTR6-1 region being largely devoid of CpG sites.

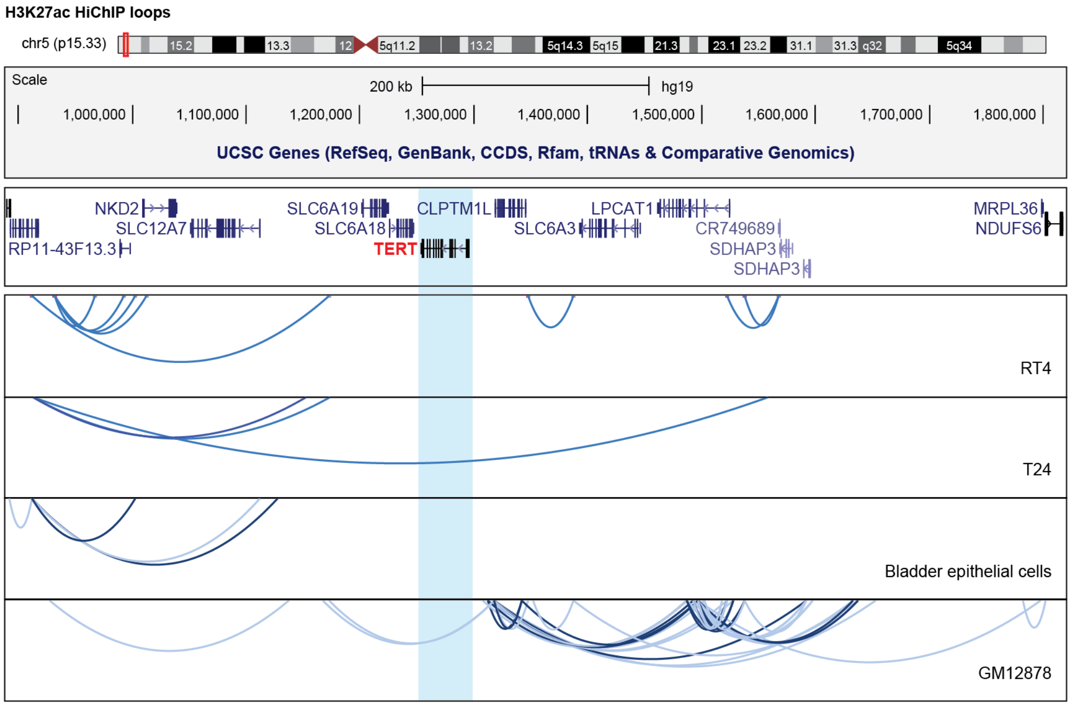


**Figure S10. Lack of long-range chromatin interactions within the *TERT* region.**

H3K27Ac HiChIP loop tracks in bladder cancer cell lines RT4 and T24 (combined biological triplicates for each cell line), normal bladder epithelial cells, and lymphoblastoid cell line GM12878. The darker-colored loops have a higher confidence based on statistical significance.

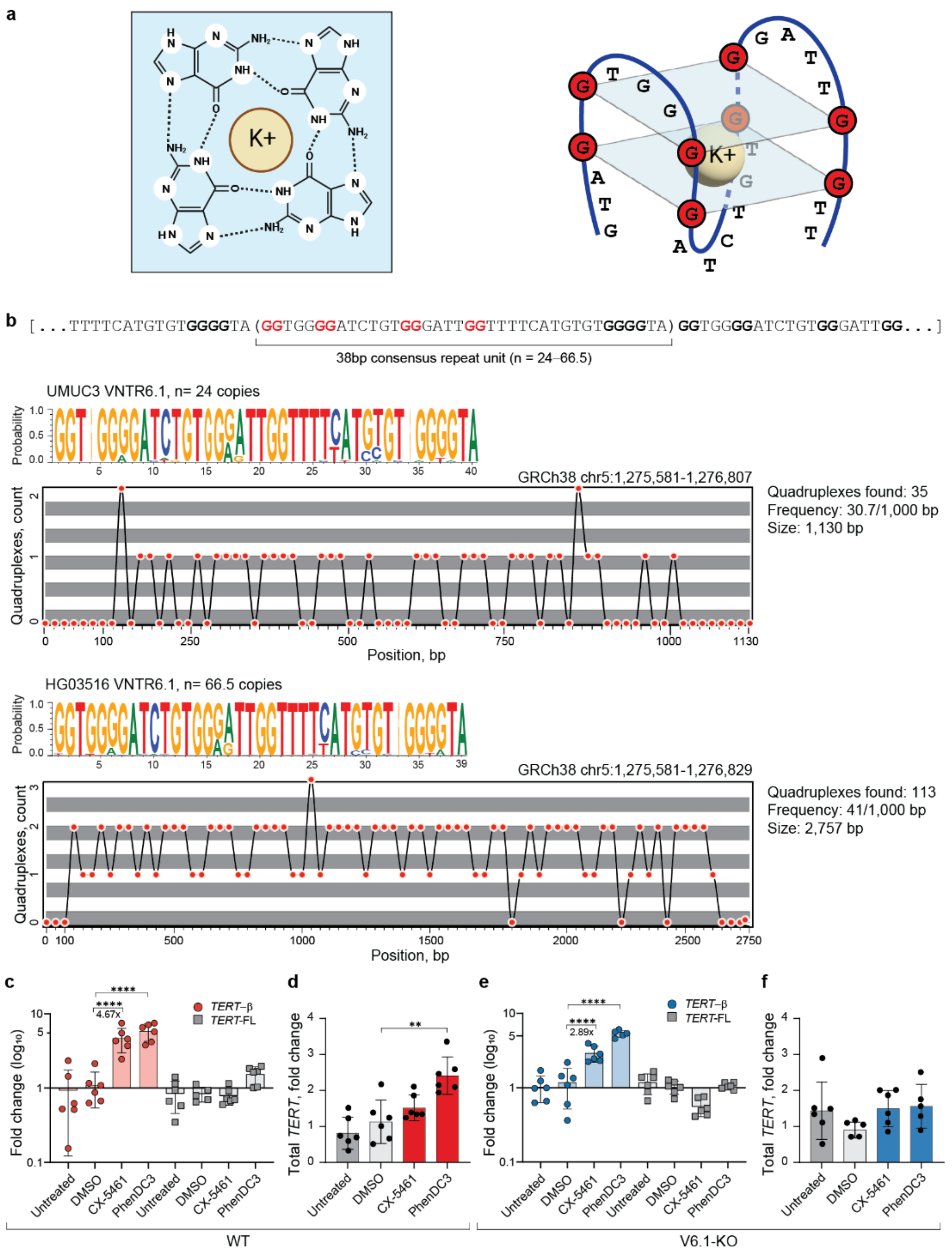


**Figure S11. Analysis of G-quadruplexes (G4) formed by VNTR6-1.**

**a**, Schematic top-view of a G4 structure characterized by hydrogen-bonded guanines (G) stabilized by a central cation (K+), created with BioRender.com; a potential model of the G4-forming folding of VNTR6-1, and VNTR6-1 consensus sequence (38 bp repeat unit) with marked guanines contributing to one G4 unit shown in the model. **b,** Sequence logos of the VNTR6-1 repeat unit and prediction of G4s across the VNTR6-1 region using G4Hunter, with each dot representing the predicted number of G4s at a corresponding position. Results are presented for one allele per sample for both the shortest VNTR6-1 version (UMUC3 cells with 24 repeat copies) and the longest VNTR6-1 version (HG03516 cells with 66.5 repeat copies) observed in human populations. **c**, **e**, Isoform-specific TaqMan RT-qPCR analysis of *TERT-FL* and *TERT-β* following treatment with G4-stabilizing ligands CX-5461 and PhenDC3 for 72 hours. **d**, **f**, Total *TERT* expression measured by TaqMan RT-qPCR assay for exon 3-4. All analyses are based on one of three representative experiments. Comparisons were made against the vehicle control (DMSO). Statistical significance is indicated as: **p < 0.01, ***p < 0.001, ****p < 0.0001, using an unpaired Student's T-test.

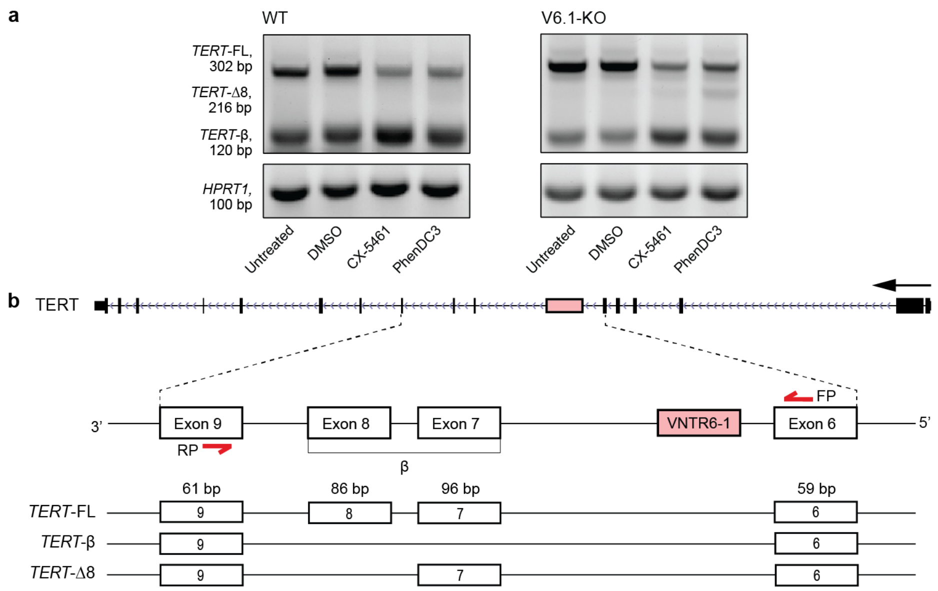


**Figure S12. Alternative splicing generates a novel *TERT*-Δ8 isoform.**

**a**, Agarose gel electrophoresis of SYBR Green qRT-PCR products from UMUC3 cells revealing a novel *TERT* isoform lacking exon 8 in cells exposed to G4 stabilizing ligands (images from **Figure 2c** and **2d**). All distinct PCR bands were gel-extracted, cloned into the TOPO-TA vector, and Sanger sequenced, confirming the identity of all PCR products and revealing that the unexpected 216-bp PCR band results from exon 8 skipping. **b**, The schematic illustrates the alternative splicing of *TERT* exons 6-9, with red arrows denoting RT-PCR primers. Exons 7 and 8 are expected to be included or skipped together; however, in the presence of G4 stabilizing ligands, exon 8 was spliced both with and without exon 7. Skipping of exon 8 (86 bp) results in a frameshift and premature transcript termination within exon 10 in a pattern similar to *TERT*-β. Both *TERT*-β and *TERT*-Δ8 produce telomerase-nonfunctional *TERT* and is likely to be eliminated by NMD.

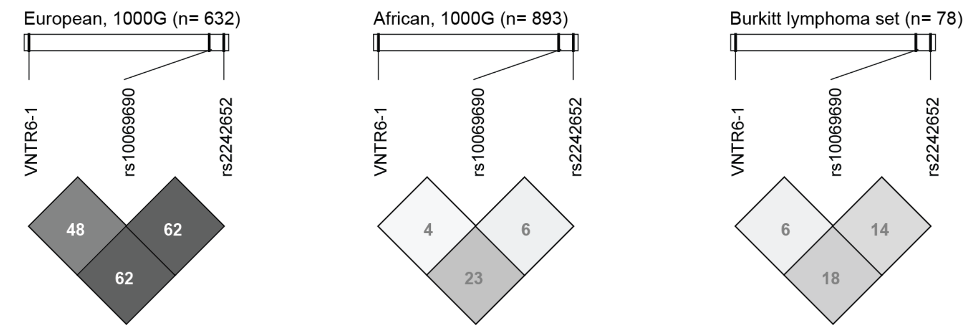


**Figure S13. Linkage disequilibrium plots ($r^2$) for individuals from 1000G-EUR and 1000G-AFR reference panels, and patients with Burkitt lymphoma (88% of African ancestry).**

The values within cells are for pairwise $r^2$ between the markers.

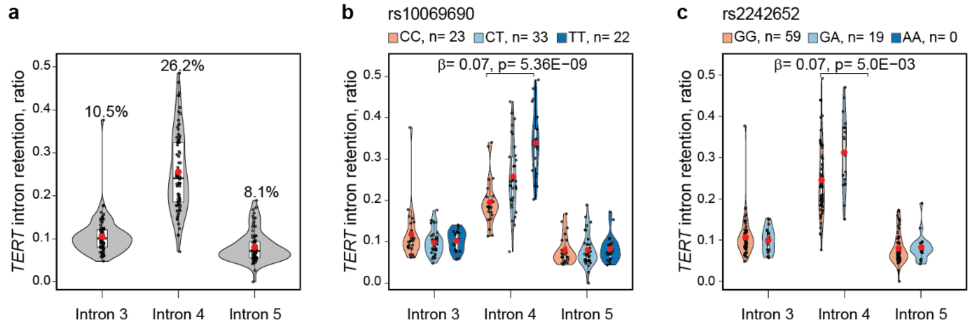


**Figure S14. Analysis of *TERT* intron retention in 78 Burkitt lymphoma tumors.**

The mRNA intron retention ratios in BL tumors were calculated for *TERT* introns 3, 4, and 5 on a scale from 0 (no retention) to 1 (full retention). **a**, Analysis in the full dataset of 79 BL tumors and in relation to genotypes of GWAS leads **b**, rs10069690 and **c**, rs2242652. The retention ratio is higher for *TERT* intron 4 than introns 3 and 5, and stronger associated with rs10069690 than rs2242652. The group means for retention ratios are shown as red dots and values above violin plots. P-values and β-values are for linear regression models adjusting for sex and age.

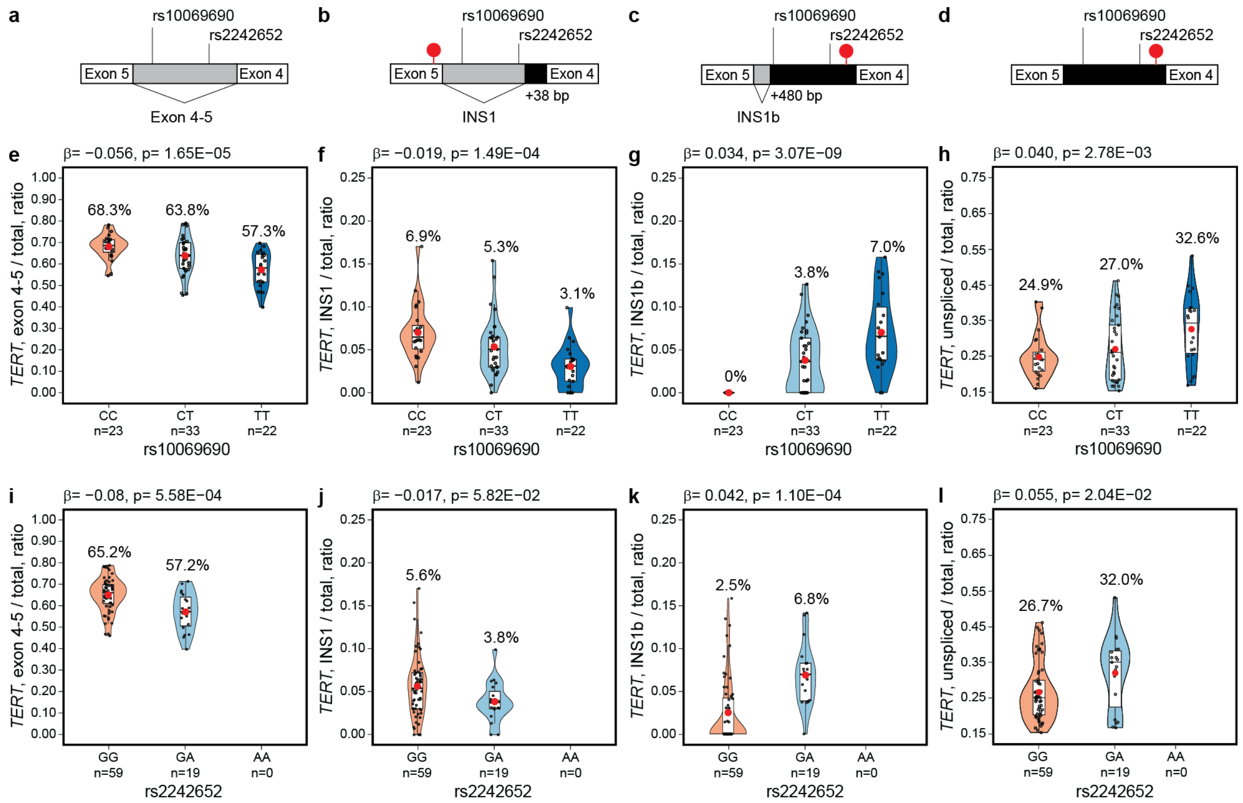


**Figure S15. Analysis of splicing events between *TERT* exons 4 and 5 in relation to GWAS leads rs10069690 and rs2242652 in 78 Burkitt lymphoma tumors.**

Schematic of *TERT* intron 4 splicing or retention, which generates **a**, canonical exon 4-5 splicing, **b**, *INS1* isoform with a stop codon in exon 5, **c**, *INS1b* isoform with a stop codon 48 bp downstream of exon 4, and **d**, unspliced intron 4 with a stop codon 48 bp downstream of exon 4. The ratios of reads and group means for rs10069690 and rs2242652 and each splicing event relative to the sum of all read counts between exons 4 and 5, including intron retention reads for **e**, **i**, canonical exon 4-5 splicing; **f**, **j**, *INS1*-type splicing; **g**, **k**, *INS1b*-type splicing; **h**, **l**, unspliced intron 4. The group means are shown as red dots and values above violin plots. White boxes – exons, gray boxes – spliced introns, black boxes – retained introns, red lollipops – stop codons. Gene direction is shown from right to left. P-values and β-values are for linear regression models adjusting for sex and age.

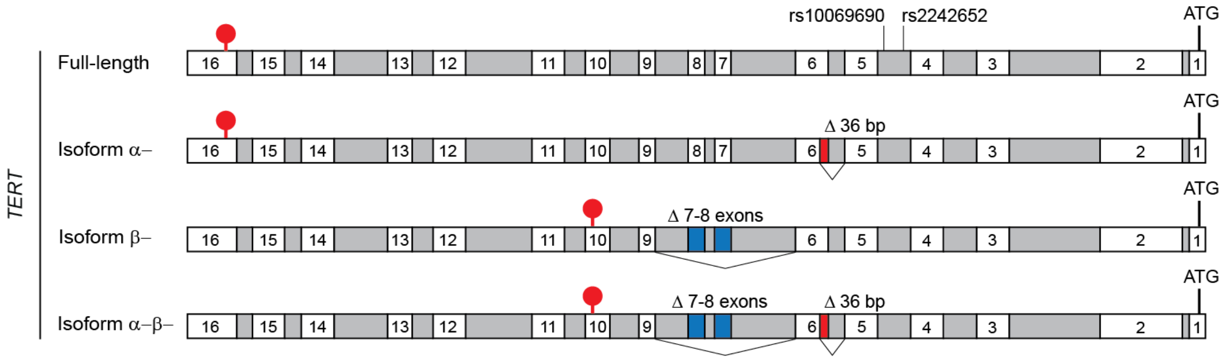


**Figure S16. Main *TERT* isoforms with alternative splicing within the area of GWAS leads rs10069690 and rs2242652.**

White boxes – constitutive exons, red boxes – alternatively spliced exons, gray boxes – introns, red lollipops – stop codons. The direction of *TERT* exons is from right to left, corresponding to the minus strand as presented in the UCSC browser. "ATG" marks translation start codons.

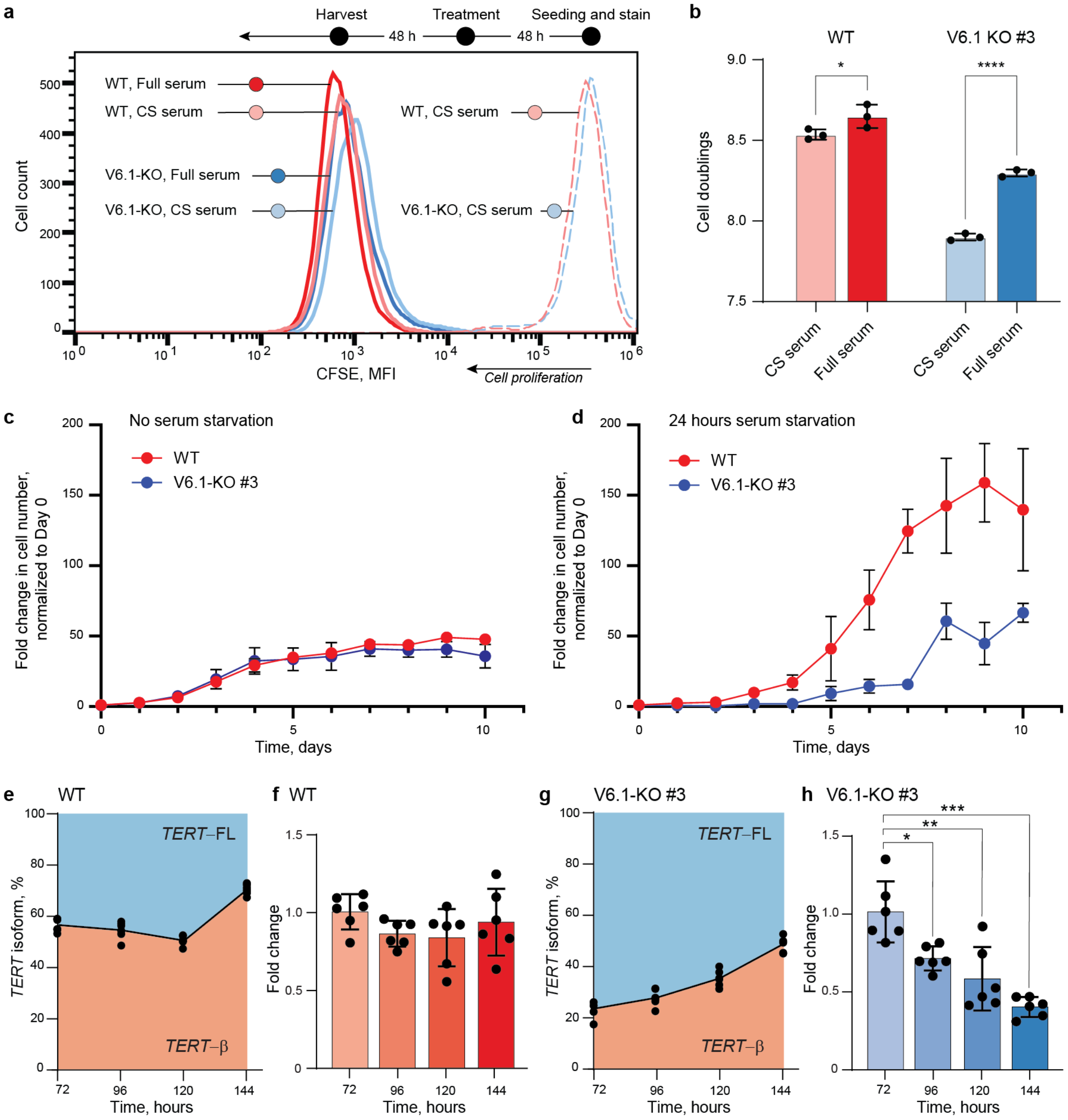


**Figure S17. VNTR6.1-associated responses to changes in environmental conditions.**

Histogram overlays of CFSE stain depletion, which occurs with every cell division, monitored over 96 hrs in both WT and VNTR6-1-KO UMUC3 cells, with the latter representing data from three different VNTR6-1-KO UMUC3 clones. The overlay plot is representative of three independent experiments (**a**). Quantification of cell doublings events in CFSE-stained cells cultured for four days in three replicates (**b**). Cell growth measured by label-free cell counting on the Lionheart automated microscope (**c**) without serum starvation and (**d**) after 24 hours of serum starvation. Average fold changes in cell counts based on four biological replicates. *TERT* isoform ratios (**e**, **g**) and total *TERT* expression (**f**, **h**) were measured over increasing time and cell density. Both analyses were based on six biological replicates, *$p < 0.05$, **$p < 0.01$, ***$p < 0.001$ Student's T-test.

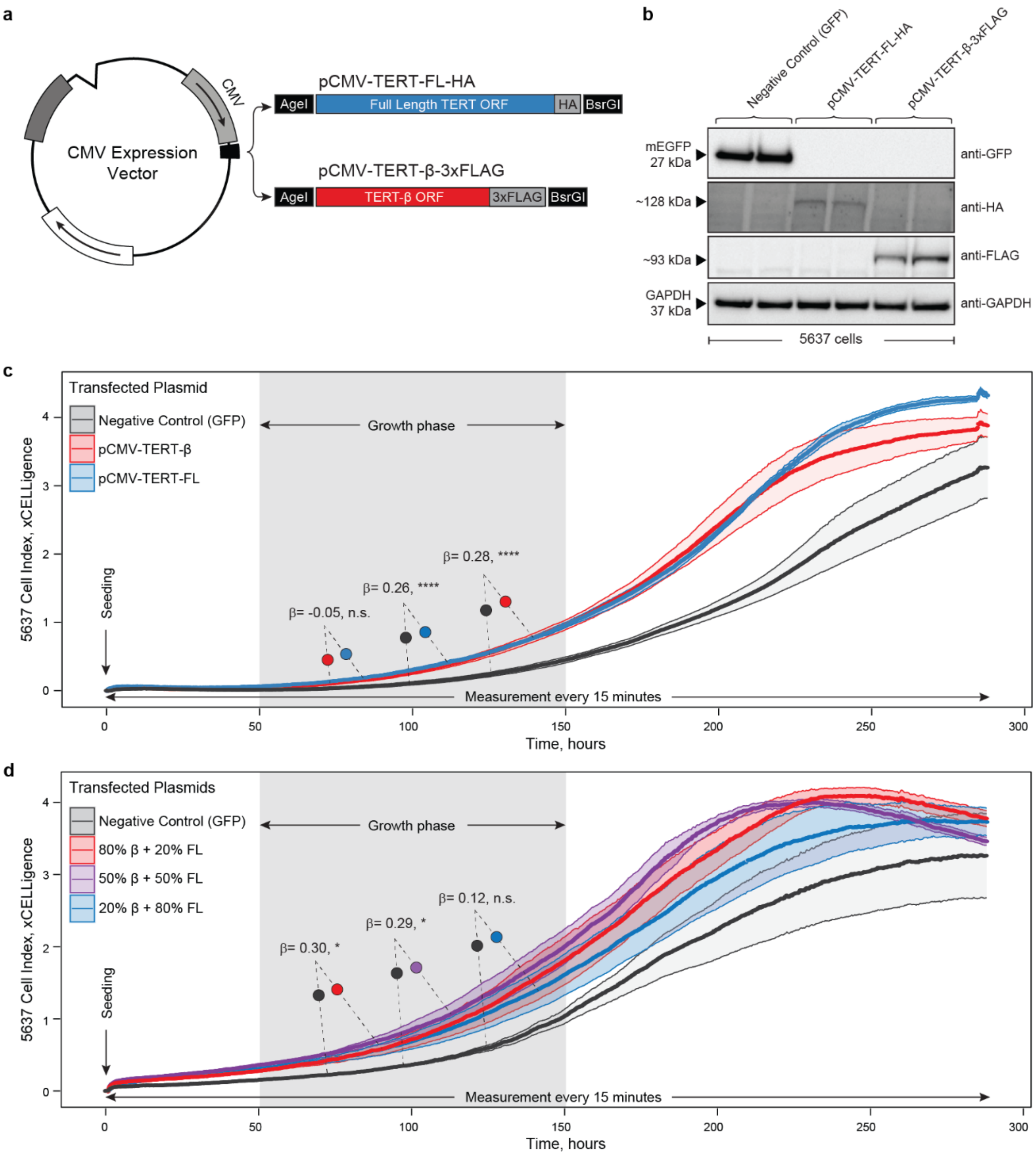


**Figure S18. Effects of overexpression of *TERT* isoforms.**

**a,** Cloning diagram of *TERT* isoforms. **b,** Western blot confirming protein levels of TERT-FL-HA and TERT-β-3xFLAG in 5637 cell line with two biological replicates each. BCA-normalized samples were used in parallel across four separate membranes which were probed with primary antibodies, including the loading control GAPDH, and processed in parallel. **c, d,** Real-time monitoring of cell growth dynamics (cell index) by measuring cellular impedance on the xCELLigence RTCA platform over 288 hours (12 days) in 5637 cells, following single transfection of *TERT* isoform expression plasmids (**c**) or co-transfection of *TERT* isoforms in various ratios of total transfected nucleic acid (TERT-FL-HA:TERT-β-3xFLAG), including 50:50, 80:20, and 20:80 (**d**). Statistical significance and β-values are for differences in cell index during the visually determined growth phase (gray highlight between 50 and 150 hrs) were calculated using linear mixed-effects models based on five (**c**) or four (**d**) technical replicates, $^{**}p < 0.01$, $^{***}p < 0.001$, $^{****}p < 0.0001$.

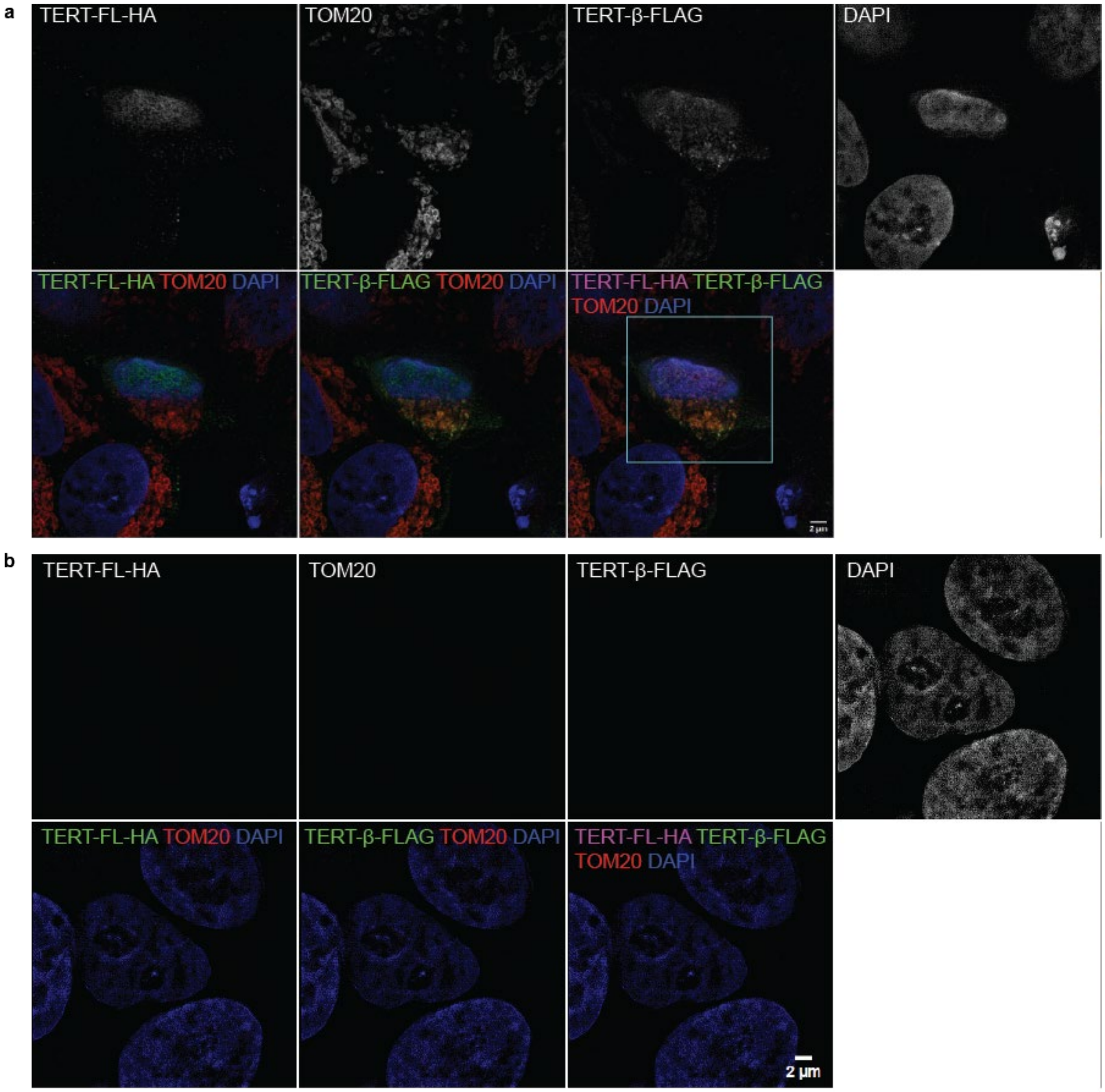


**Figure S19. Cellular localization of the TERT-FL and TERT-β protein isoforms.**

Structured illumination microscopy (SIM) images of the TERT-FL and TERT-β protein isoforms transiently overexpressed in the A549 lung cancer cell line. Cells were co-transfected with TERT-FL-HA and TERT-β-FLAG expression constructs at a 50:50% ratio and stained with corresponding antibodies. For individual channels, staining is shown as black/white images for better contrast. On tri-color merged panels, green – FLAG (TERT-β) or HA (TERT-FL), blue – DAPI (nuclei). On the quad-color merged panel, purple – HA (TERT-FL), green – FLAG (TERT-β), red – TOM20 (mitochondria), blue – DAPI (nuclei). **a**, zoomed-out view of transfected cells stained with anti-FLAG and anti-TOM20 primary antibodies, as well as respective secondary antibodies AlexaFluor647-/AlexaFluor488- (pseudocolored green) and AlexaFluor555- (pseudocolored red). The orange color indicates the mitochondrial colocalization of TOM20 with TERT-β, not seen for TERT-FL; **b**, zoomed-out view of negative controls: similarly transfected cells but omitting primary antibodies and stained only with secondary antibodies, showing signals are not from autofluorescence. Confocal images were captured with a 63x/1.4 NA objective. Scale bar = 2 µm, except for the inset panel (0.5 µm).

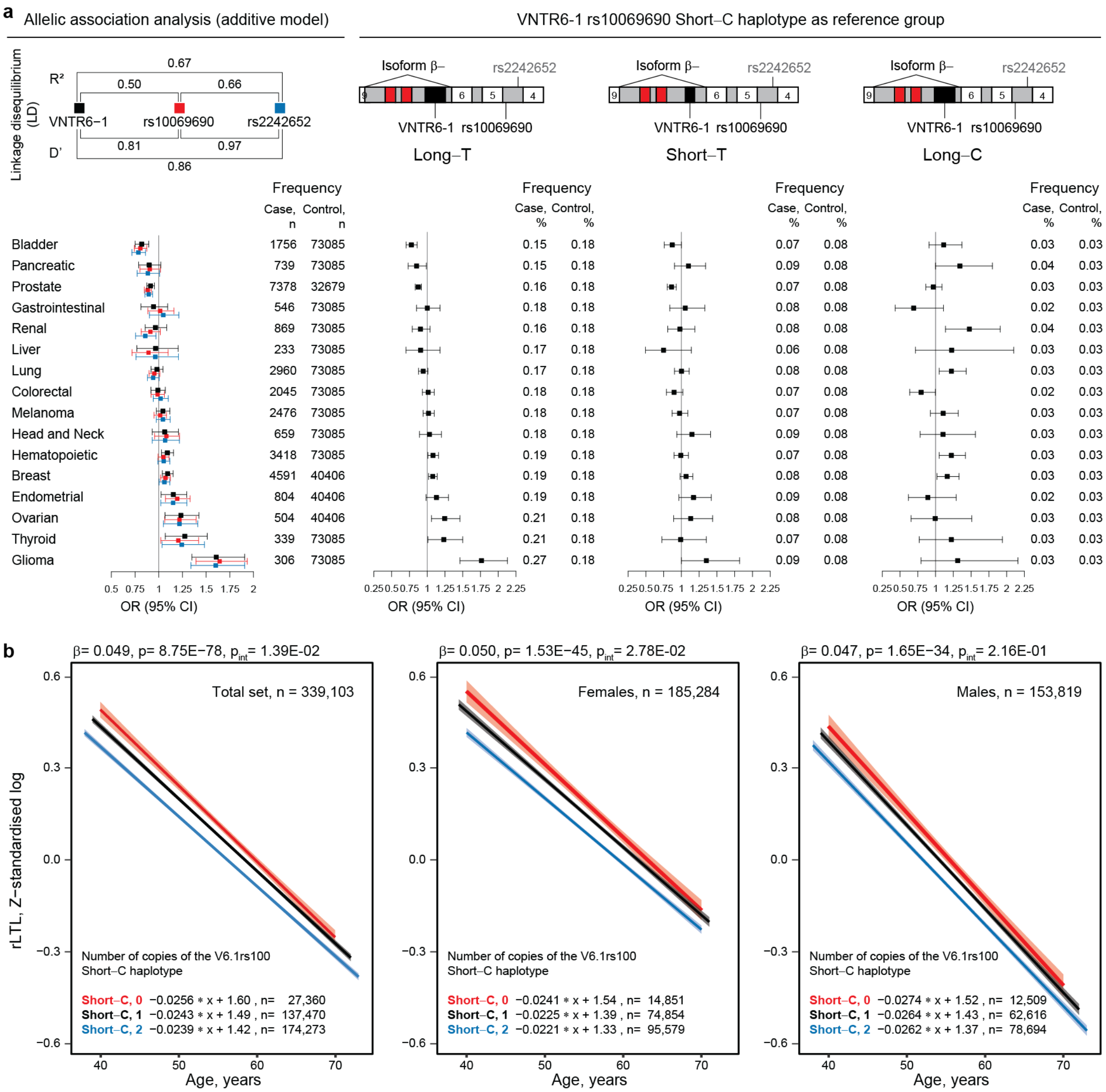


**Figure S20. Association analysis of cancer risk in PLCO and relative leukocyte telomere length (rLTL) in UKB cancer-free individuals.**

**a,** Evaluation of cancer risk associated with VNTR6-1-Long, rs10069690-T and rs2242652-A alleles and haplotypes of the composite marker (VNTR6-1/rs10069690) compared to the Short-C haplotype) in the PLCO dataset (n=102,708). Odds Ratios (OR) with 95% confidence intervals (CI) were calculated comparing patients with indicated cancers to a common group of cancer-free controls, using logistic regression analysis with an additive genetic model, adjusting for sex and age. **b**, Evaluation of the relationship between rLTL and the VNTR6-1/rs10069690 marker in UKB cancer-free individuals (n=339,103). P-values and β-values were derived from linear regression models, adjusted for sex, age, and smoking status, and for age and smoking status in sex-specific analyses (**Table S15**). $P_{int}$ are for interaction between genotypes and age groups. The graphs display regression lines with 95% confidence intervals and regression equations. The analysis shows a decrease in rLTLs with more copies of the Short-C haplotype.

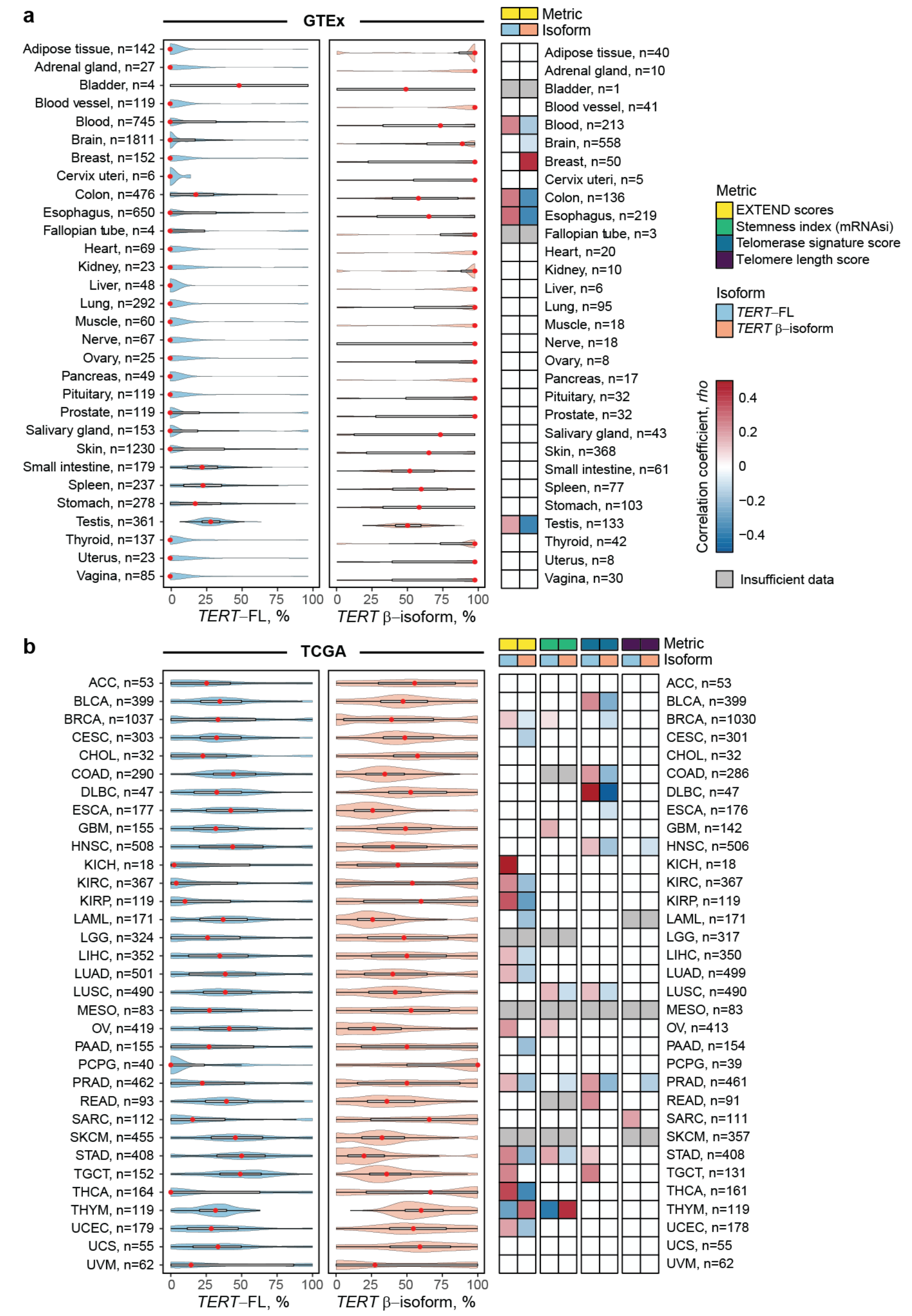


**Figure S21. Analysis of isoform-level *TERT* expression in relation to telomerase-associated metrics in GTEx and TCGA.**

Isoform-level *TERT* expression (full-length and β-isoform) as a percentage of total *TERT* expression analyzed as transcripts per million (TPMs) in **a**, normal tissue samples in GTEx and **b**, tumors in TCGA. The mean expression for each tissue and tumor type is represented by a red dot within each violin plot. Spearman rank correlation coefficients (rho) between *TERT* expression (full-length and β-isoform) and various telomerase-associated metrics in **a**, GTEx and **b**, TCGA. Only statistically significant correlations are shown in color, gray – insufficient data for analysis; n indicates the number of samples analyzed. TCGA data is for one sample per individual, GTEx data combined samples from each tissue type, for some tissues (brain, skin) this includes several samples per individual. Full results are presented in **Table S14**.

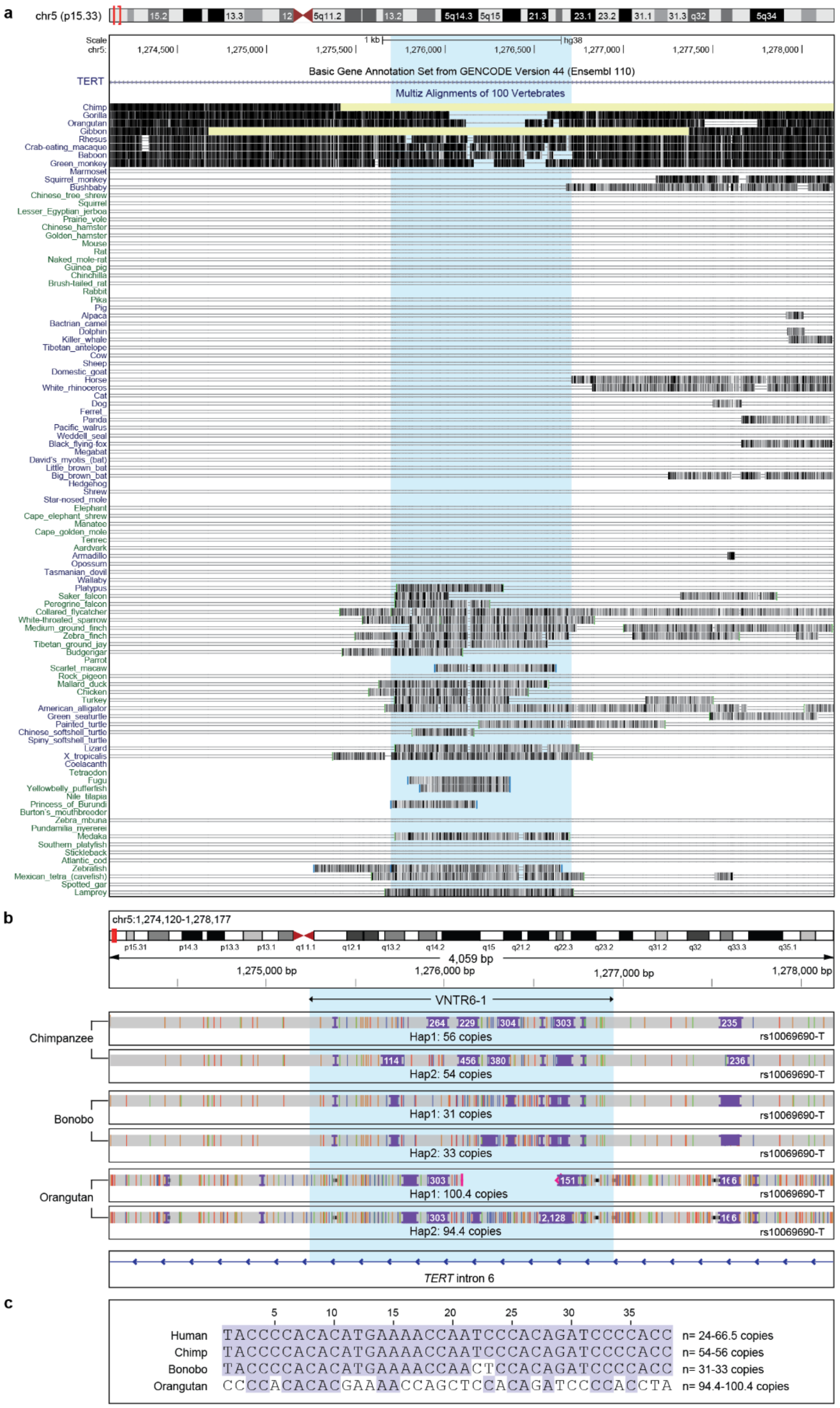


**Figure S22. *TERT* VNTR6.1 region across vertebrate species.**

**a,** Multiple sequence alignment in 100 vertebrate species (source: UCSC Genome Browser). **b,** IGV plots of primate Telomere-to-Telomere assemblies aligned to the human genome (GRCh38). Shown are the VNTR6-1 region (blue highlight) with the number of repeat copies per haplotype and rs10069690 alleles within the same haplotype. **c,** Comparison of the VNTR6-1 consensus repeat unit (38 bp) between humans and primates.

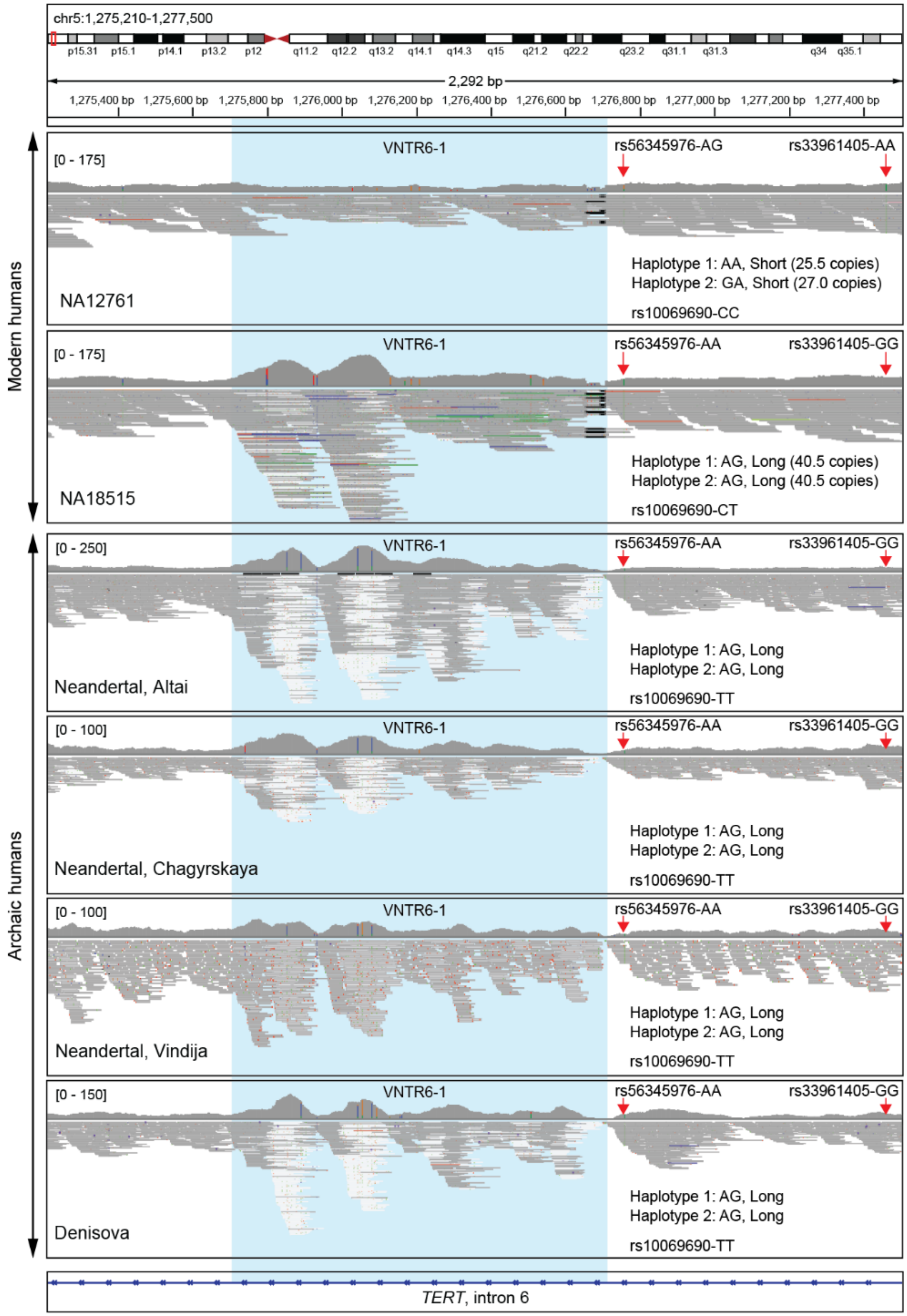


**Figure S23.** ***TERT*** **VNTR6-1 profiles in WGS short-read alignments in modern and archaic humans.**

IGV plots of WGS short-read sequencing (Illumina, 30x coverage) illustrating VNTR6-1 (blue highlight) and SNPs rs56345976 and rs33961405 in modern humans (NA12761, Short/Short, and NA18515, Long/Long) and archaic humans (Neandertal and Denisova, all VNTR6-1-Long/Long genotypes).

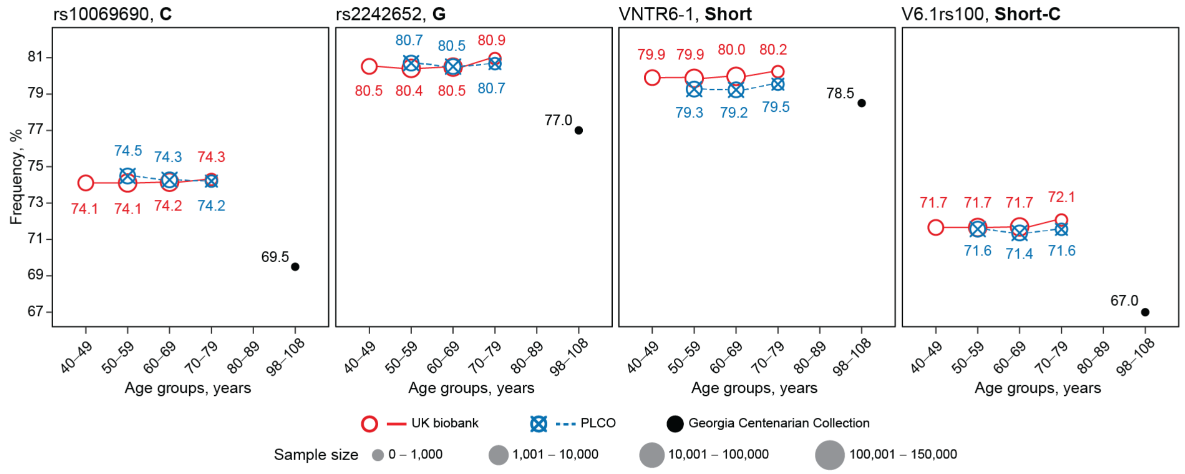


**Figure S24. Age-dependent frequencies of the human-specific alleles in individuals of European ancestry.**

The distribution of the derived alleles of markers in cancer-free individuals from UK biobank (n= 351,629) and PLCO (n= 73,084), and in the Georgia Centenarian Collection (n= 100). Dot sizes correspond to the sample numbers in each age group. Alleles rs10069690-C and VNTR1-Short and Short-C haplotype are associated with higher total *TERT* expression and a lower ratio of alternative non-telomerase producing isoforms *INS1b* (rs10069690-C) and *TERT-b* (Short). In this set of centenarians of European ancestry, the decrease in frequencies of derived alleles is unlikely due to potential population stratification, as the frequency of the derived rs2242652-G allele is the lowest in the populations of European ancestry (1000G-EUR, 79%), while higher in other populations (1000G-AFR, 88%, 1000G-EAS, 85%, 1000G-AMR, 85%, 1000G-SAS, 82%). A potential admixture with other populations would result in higher, not lower rs2242652-G allele frequency observed in centenarians of European ancestry (77.0%).
